# Interindividual HLA Evolutionary Divergence in Single HLA-Mismatched Unrelated Donor Hematopoietic Cell Transplantation for Malignant Hematological Disorders: A Report on Behalf of the Cellular Therapy and Immunobiology Working Party of the EBMT

**DOI:** 10.64898/2026.02.22.26346823

**Authors:** Simona Pagliuca, Jarl E. Mooyaart, Francis Ayuk, Robert Zeiser, Victoria Potter, Peter Dreger, Wolfgang Bethge, Inken Hilgendorf, David Michonneau, Alessandro Rambaldi, Henrik Sengeloev, Jakob Passweg, Deborah Richardson, Tobias Gedde-Dahl, Francesca Kinsella, Matthias Edinger, Stephan Mielke, Matthias Eder, Marco Andreani, Pietro Crivello, Pietro Merli, Jorinde D. Hoogenboom, Liesbeth C. de Wreede, Christian Chabannon, Jürgen Kuball, Carmelo Gurnari, Katharina Fleischhauer, Annalisa Ruggeri, Tobias L. Lenz

**Affiliations:** Service d’hématologie Clinique, Hôpital Brabois, CHRU Nancy and CNRS UMR 7365 IMoPa, Biopole de l’Université de Lorraine, Nancy, France; EBMT Leiden Study Unit, Leiden, The Netherlands; University Hospital Eppendorf, Hamburg, Germany; Faculty of Medicine, Clinic for Internal Medicine I, Hematology, Oncology and Stem Cell Transplantation, Faculty of Medicine, University Medical Center Freiburg, Freiburg, Germany; Kings College Hospital London, London, United Kingdom; University of Heidelberg, Heidelberg, Germany; University Hospital Tuebingen, Department of Hematology, Oncology, Clinical Immunology and Rheumatology, Tuebingen, Germany; Universitaetsklinikum Jena, Jena, Germany; Hematology and transplantation unit, Saint Louis hospital, APHP, Paris, France; INSERM U1342, Saint Louis Research Institute, Paris Cité University, Paris, France; ASST Papa Giovanni XXIII, Bergamo, Italy; Rigshospitalet, Copenhagen, Denmark; University Hospital | Basel, Basel, Switzerland; Southampton General Hospital, Southampton, United Kingdom; Oslo University Hospital, Rikshospitalet, Oslo, Norway; Birmingham Centre for Cellular Therapy and Transplant (BCCTT), Birmingham, United Kingdom; University Regensburg, Regensburg, Germany; Karolinska University Hospital, Stockholm, Sweden; Department of Hematology, Hemostasis, Oncology and Stem Cell Transplantation, Hannover Medical School; IRRCS Ospedale Pediatrico Bambino Gesù, Rome, Italy; Institute for Experimental Cellular Therapy, University Hospital Essen, Essen, Germany; Department of Biomedical Data Sciences, LUMC, Leiden, The Netherlands; Institut Paoli-Calmettes Comprehensive Cancer Centre and Module Biothérapies du Centre d’Investigations Cliniques de Marseille, Marseille, France; University Medical Center Utrecht, Utrecht, The Netherlands; Department of Biomedicine and Prevention, University of Rome Tor Vergata, Rome, Italy; San Raffaele Scientific Institute, Hematology and Bone Marrow Transplantation Unit, Milan, Italy; Research Unit for Evolutionary Immunogenomics, Department of Biology, University of Hamburg, Hamburg, Germany

## Abstract

Allogeneic hematopoietic cell transplantation (allo-HCT) hinges on a delicate trade-off between graft-versus-tumor control and graft-versus-host disease (GvHD), mediated by donor T-cell recognition of antigens presented by recipient human leukocyte antigen (HLA) molecules. We hypothesized that, beyond allele-level matching, sequence divergence at peptide-binding grooves across donor and recipient HLA loci shapes these responses. To this end, we evaluated the effect of HLA evolutionary divergence (HED), a metric quantifying amino acid variability at HLA peptide-binding sites, on selected hematological malignancies in 4,695 patients undergoing allo-HCT from a 9/10 mismatched unrelated donor (MMUD), reported to the EBMT database. We examined (i) locus-specific recipient HED (HED-R) and (ii) “HED-mismatch” (HED-MM), capturing immunopeptidome divergence at the mismatched locus. While dichotomous mismatch status explained differences in survival and acute GvHD risk (with overall greater detriment for class I loci), HED metrics uncovered substantial within-mismatch heterogeneity. In DRB1 mismatched subgroup, HED-MM at this locus, independently predicted inferior relapse-free survival (RFS) with an attenuating time-dependent association, further modulated by cross-locus HED-R. In this subgroup, higher HED-R at HLA-A and HLA-C associated with increased risks of acute GvHD and non-relapse mortality, respectively. Among HLA-B–mismatched pairs, higher DRB1 HED-R associated with worse overall survival (OS) and RFS and higher relapse risk. In the HLA-A–mismatched subgroup, higher HED-R at HLA-A increased chronic GvHD risk. Collectively, HED-derived metrics complement conventional mismatch classification by capturing qualitative differences in donor–recipient immunopeptidome interactions and reveal a complex, non-linear interplay among alleles across mismatch subgroups that modulates the clinical impact of mismatching.

**Keypoints:**

- In mismatched unrelated HCT, baseline risk varies across mismatch constellations, with class I mismatches more detrimental than class II.
- HED complements conventional HLA mismatch classification by capturing qualitative donor-recipient immunopeptidome interactions.

## Introduction

Allogeneic hematopoietic cell transplantation (allo-HCT) remains a curative strategy for many hematologic malignancies, yet outcomes are constrained by the narrow therapeutic window between graft-versus-tumor (GvT) activity and graft-versus-host disease (GvHD) ^1–3^. Both processes are initiated by donor T-cell recognition of recipient-derived peptides presented by human leukocyte antigen (HLA) molecules. Consequently, donor–recipient HLA disparities are central determinants of post-transplant alloreactivity and clinical outcome^4,5^.

Current unrelated donor selection primarily relies on allele-level HLA matching and other non-HLA factors, and transplantation from a single-mismatched unrelated donor (9/10 MMUD) is often required when a fully matched donor or another suitable related donor is unavailable. However, the clinical impact of HLA mismatching is not uniform with differences by locus being well recognized, and substantial heterogeneity existing within the same mismatch category^5^. Indeed, the mere allele-level mismatches incompletely capture the functional consequences of donor–recipient HLA disparity^5–8^. In this context, several approaches have moved beyond a sole quantitative approach by incorporation of the quality of the mismatch. Notably, the HLA-DPB1 T-cell epitope (TCE) model distinguishes permissive from non-permissive mismatches and has been shown to reflect biologically meaningful differences in alloreactivity, consistent with differential immunopeptidome divergence between donor and recipient at locus DPB1^9^. Complementing this concept, recent work has proposed evaluating donor–recipient divergence at the level of peptide-binding motifs (PBM), differences in the anchor-residue preferences that shape which peptides are presented by a given HLA molecule. PBM divergence frameworks aim to operationalize “functional distance” between alleles by approximating the difference between peptide repertoires of patients and donors, allowing the identification of clinically relevant HLA disparities among seemingly comparable mismatch categories.^10,11^ Together, these immunopeptidome-oriented models support the broader premise that donor selection and risk stratification may benefit from metrics that approximate how much peptide presentation differs between donor and recipient, introducing a certain degree of heterogeneity in terms of functional immunopeptidome presentation, rather than treating all allele mismatches as equivalent.

Recently, aligning with these approaches, the concept of HLA evolutionary divergence (HED), a metric quantifying the amino acid difference between two paired HLA alleles, has been proposed as a proxy for assessing an individual’s immunocompetence^12,13^. Because the peptide-binding groove largely determines which peptides can be presented, this metric provides a structured, sequence-based approximation of potential immunopeptidome diversity, with greater sequence divergence at these sites expected to translate into more distinct peptide repertoires. This parameter has demonstrated its relevance across various settings, from susceptibility to infections and autoimmunity to responses to immunotherapy in patients with solid cancers^12–15^. We, along with others, have extended its application to the field of allo-HCT, utilizing HED to evaluate the propensity for alloreactivity^16–19^. We and others observed that in a matched setting, recipient HED at specific loci may influence the risk of post-transplant outcomes in both adult and pediatric settings and may correlate with the diversity of T-cell receptor (TCR) repertoire and other markers of immune reconstitution^16–18^. Indeed, while individual HED values (for donor and recipient) may already carry important immunological implications, the next frontier involves examining how interindividual HED differences might translate into donor-recipient immunopeptidome disparities at a mismatched locus.

Here, we investigated whether HED-based metrics refine outcome prediction in MMUD allo-HCT beyond conventional locus-based mismatch classification. Specifically, we assessed (i) single-locus donor and recipient HED (HED-D and HED-R) and (ii) an interindividual “HED-mismatch” metric (HED-MM), defined as the immunopeptidome divergence attributable to the mismatched donor– recipient allele pair at a given locus. Using a large EBMT cohort of patients receiving 9/10 MMUD allo-HCT for hematologic malignancies, without post-transplant cyclophosphamide (PTCy), we examined associations of these metrics with survival and major transplant endpoints, testing whether immunopeptidome-oriented HED measures capture clinically relevant heterogeneity within traditional mismatch groups and thereby improve risk stratification after allo-HCT.

## Methods

### Study design and cohort

This retrospective registry-based study used multicenter data from the European Society for Blood and Marrow Transplantation (EBMT) to investigate the impact of HED on outcomes after allogeneic hematopoietic cell transplantation (allo-HCT) from single mismatched unrelated donors (9/10 MMUD).

Eligible patients were adults (≥18 years) with acute myeloid leukemia, acute lymphoblastic leukemia, myelodysplastic syndromes, or myeloproliferative neoplasms who underwent a first allo-HCT from a 9/10 MMUD between January 2010 and July 2019 at EBMT-affiliated centers. Both myeloablative and reduced-intensity conditioning regimens were included. To isolate immunogenetic effects of HLA mismatching, patients receiving post-transplant cyclophosphamide (PT-Cy) were excluded. Graft sources were bone marrow or peripheral blood.

Only patients with high-resolution (2-field) HLA typing available for loci A, B, C, DRB1, and DQB1 were included. A subset of patients with DPB1 mismatches was analyzed separately. The study was conducted in accordance with the Declaration of Helsinki. Data were collected within the EBMT Registry after patients provided written, GDPR (General Data Protection Regulation)**-**compliant informed consent for data registration and research use, according to EBMT Registry governance; participating centres complied with applicable national/institutional requirements for registry reporting.

### HED metrics

HED metrics were computed using a validated sequence-based algorithm based on Grantham distance, quantifying amino acid divergence at peptide-binding regions of HLA molecules^13^ (**Figure 1A**).

**Figure 1.**
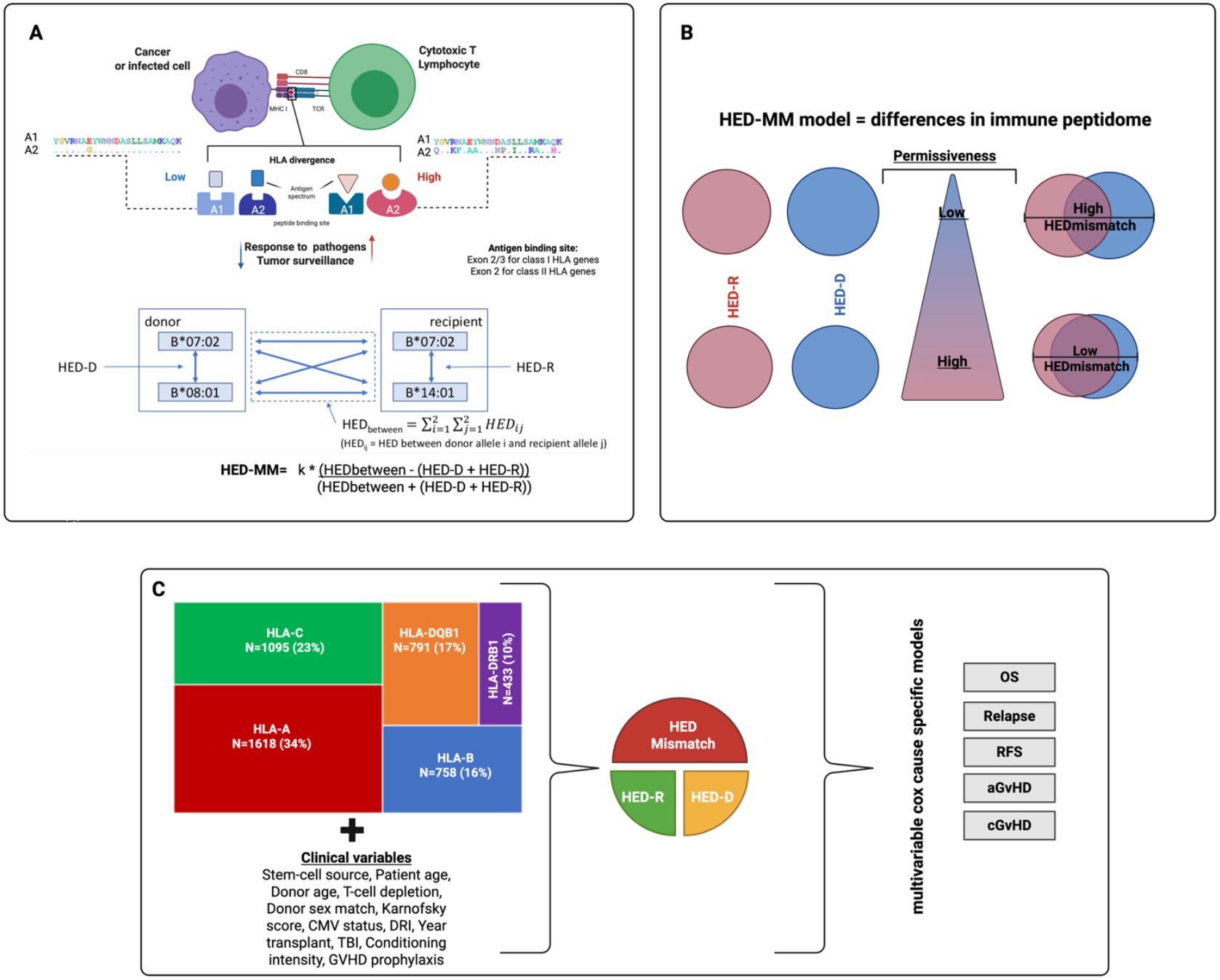
Conceptual framework and study design for assessing the impact of HLA evolutionary divergence (HED) in mismatched unrelated donor (MMUD) allo-HCT. **(A)** *HLA evolutionary divergence (HED) and computation of HED mismatch*. HED quantifies the amino-acid dissimilarity between the two alleles carried at a given HLA locus, focusing on the antigen-binding site (exons 2–3 for class I; exon 2 for class II). Higher within-individual HED is expected to broaden the immunopeptidome presented to T cells. HED metrics are computed by measuring amino acid variability within HLA molecules and between donor-recipient pairs. A1 and A2 indicate two generic alleles within the same locus. Donor (HED-D) and recipient (HED-R) within-locus divergence are combined (HED_between; pairwise distances between each donor allele and each recipient allele) to derive an HED mismatch score (HEDmismatch, HED-MM), reflecting the extent to which donor and recipient differ in their presented peptide landscapes. **(B)** *HED-MM model and permissiveness*. Schematic linking donor–recipient immunopeptidome divergence to mismatch permissiveness using HED-MM. Donor (blue) and recipient (pink) locus-specific HED profiles are combined to estimate the extent to which their antigen-presenting landscapes differ. High HED-MM reflects greater divergence (reduced overlap between donor and recipient peptide-presenting repertoires) and is therefore expected to correspond to a less permissive mismatch, whereas low HED-MM indicates more similar donor and recipient HED profiles (greater overlap) and a more permissive mismatch. The right-hand examples illustrate these two extremes. **(C)** *Analytical workflow*. Distribution of locus-specific mismatch groups included in the analyses (HLA-A, -B, -C, -DRB1, -DQB1; sample sizes shown). For each mismatched locus, models incorporated HED-derived metrics (HED-R, HED-D and HED-MM) alongside key clinical covariates (stem-cell source, recipient/donor age, T-cell depletion, donor sex match, Karnofsky score, CMV status, DRL, transplant year, TBI, conditioning intensity, GvHD prophylaxis) in multivariable cause-specific Cox models for overall survival (OS), relapse, relapse-free survival (RFS), acute GvHD (grade II–IV), and chronic GvHD.

Two categories of HED metrics were evaluated (**Figure 1B**):

- Individual HED, calculated at each locus: Recipient HED (HED-R): divergence between the recipient’s two alleles. Donor HED (HED-D): divergence between the donor’s two alleles.
- Interindividual HED-mismatch (HED-MM), computed only at the mismatched locus, captures the functional distance introduced by the donor–recipient HLA disparity by quantifying amino-acid divergence between the non-shared donor and recipient alleles at that locus. HED-MM thus provides a locus-specific proxy of mismatch-driven immunopeptidome dissimilarity, with higher values indicating more divergent peptide-binding architectures. Extended methodological details and biological rational of HED metrics are provided in the *Supplementary Methods*.

### Statistical analysis

Baseline characteristics were summarized using descriptive statistics.

The primary endpoints were overall survival (OS) and relapse-free survival (RFS). Secondary endpoints included the cumulative incidence of relapse, non-relapse mortality (NRM), acute graft-versus-host disease (grade II–IV), and chronic graft-versus-host disease, taking into account competing risks (see Supplementary Methods).

OS and RFS of subgroups were compared using Kaplan–Meier estimates and log-rank tests. Cumulative incidence functions were used for endpoints with competing risks and compared using Gray’s test.

Associations between HED metrics and outcomes were assessed using (cause-specific) Cox proportional hazards models. Each HED metric was evaluated in separate multivariable models, adjusted for clinically relevant covariates, including diagnosis, modified disease risk index (DRI)^220^, year of transplant, patient and donor age, CMV serostatus, conditioning intensity, graft source, T-cell depletion, GvHD prophylaxis, total body irradiation, donor–recipient sex mismatch, and Karnofsky performance status. The proportional hazards assumption was tested, and time-dependent associations were modeled when it was violated. All analyses were performed within locus-specific mismatch subgroups. Two-sided p-values <0.05 were considered statistically significant. Further statistical details are provided in the *Supplementary Methods*.

## Results

### Overview of the study cohort and metrics description

Overall, 4695 adult patients, with diagnoses of acute leukemia (N=3897, 83%) and myelodysplastic syndrome or myeloproliferative neoplasms (N=798, 17%) were identified **(Table 1)**. Median recipient age was 53.5 (40.2-62.0) with a male/female ratio of 1.29, whilst median donor age was 31.3 (24.9-40). Majority of patients (91%) received peripheral blood stem cells and were mismatched with their donors for locus A (N=1618 34%), followed by C (N=1095, 23%), B (N=758, 16%), DQB1 (N=791, 17%) and DRB1 (N=433, 10%). Among them, and for patients with available information, N= 1389 had a DPB1 mismatch across all the loci. The most frequent donor-recipient sex pairings were male-to-male (41%) and female-to-male (25%), while male-to-female (15%) and female-to-female (19%) transplants were less common. The study spanned a decade, with nearly half of the transplants (48%) performed between 2010 and 2014, and the remaining 52% between 2015 and 2019. Disease risk at transplant, classified by the Disease Risk Index (DRI), showed that 31% of patients had a low-to-intermediate risk, while 69% had a high or very high risk. Concerning T-cell depletion strategies, 84% of patients received in vivo T-cell depletion, while the remaining underwent either ex vivo depletion or a combination of both in vivo and ex vivo approaches. In terms of conditioning regimen, 49% of patients received standard-intensity conditioning and 51% reduced-intensity regimens. GvHD prophylaxis strategies varied, with ciclosporin and methotrexate (CSA-MTX) being the most frequently used regimen (44%), followed by ciclosporin and mycophenolate mofetil (CSA-MMF) (27%), Tacrolimus-based regimens (12%), and other combinations (16%). Total body irradiation (TBI) was employed in 25% of cases. Distribution of HED metrics followed locus-specific patterns (**Figure S1A**) and scores related to HED-R were not affected by disease categories (**Figure S1B**).

**Table 1.**

| Patient characteristics N (%) |  |
| --- | --- |
| All | 4,695 |
| **Patient sex** |  |
| Male | 2,646 (56%) |
| Female | 2,049 (44%) |
| **Stem-cell source** |  |
| BM | 446 (9%) |
| PB | 4,249 (91%) |
| **Patient age** |  |
| >=55 | 2,168 (47%) |
| 18-35 | 866 (18%) |
| 35-55 | 1,661 (35%) |
| **Donor age** |  |
| >=55 | 61 (1%) |
| 18-35 | 2,113 (45%) |
| 35-55 | 1,228 (26%) |
| missing | 1,293 (27%) |
| **Disease type** |  |
| Acute Leukemia | 3,897 (83%) |
| SMD/MPN | 798 (17%) |
| **Year transplant** |  |
| 2010-2014 | 2,275 (48%) |
| 2015-2019 | 2,420 (52%) |
| **DRI** |  |
| Intermediate-Low | 1,473 (31%) |
| High-Very High | 3,222 (69%) |
| **Karnofsky score** |  |
| >=90 | 3,263 (69%) |
| <90 | 1,133 (24%) |
| missing | 299 (7%) |
| **Donor sex match** |  |
| recipient male - donor male | 1,914 (41%) |
| recipient male - donor female | 705 (15%) |
| recipient female - donor male | 1,138 (24%) |
| recipient female - donor female | 893 (19%) |
| Missing (NA) | 45 (1%) |
| **CMV status (Donor/Recipient)** |  |
| -/- | 1,093 (24%) |
| -/+ | 434 (9%) |
| +/- | 1,363 (29%) |
| +/+ | 1,665 (35%) |
| Missing (NA) | 140 (3%) |
| **T-cell depletion** |  |
| no | 585 (12%) |
| yes invivo, no exvivo | 3,953 (84%) |
| yes exvivo, no invivo | 10 (0.3%) |
| yes invivo+exvivo | 73 (1.8%) |
| missing exvivo, no invivo | 10 (0.3%) |
| missing exvivo, yes invivo | 64 (1.6%) |
| **Conditioning intensity** |  |
| standard | 2,273 (48.4%) |
| reduced | 2,390 (51%) |
| Missing (NA) | 32 (0.6%) |
| **GVHD prophylaxis** |  |
| CSA-MTX | 2,058 (44%) |
| CSA_MMF | 1,284 (27%) |
| Tac-based | 580 (12%) |
| Other | 773 (17%) |
| **Total body irradiation** |  |
| Yes | 1,173 (25%) |
| Missing (NA) | 10 |
| **ABO patient** |  |
| A | 796 (17%) |
| B | 177 (3.8%) |
| AB | 88 (1.9%) |
| O | 720 (15,3%) |
| unknown | 2,914 (62%) |
| **HLA Mismatch** |  |
| Mismatch A | 1,618 (34%) |
| Mismatch B | 758 (16%) |
| Mismatch C | 1,095 (23%) |
| Mismatch DQB1 | 791 (17%) |
| Mismatch DRB1 | 433 (10%) |
| **Nr of mismatch DPB1** |  |
| 0 | 277 (5.9%) |
| 1 | 890 (19%) |
| 2 | 499 (11%) |

**Abbreviations:** ABO, ABO blood group; BM, bone marrow; CMV, cytomegalovirus; CSA, cyclosporine A; DRI, Disease Risk Index; GVHD, graft-versus-host disease; HCT, hematopoietic cell transplantation; HLA, human leukocyte antigen; MMF, mycophenolate mofetil; MTX, methotrexate; NA, not available; PB, peripheral blood; Tac, tacrolimus; TBI, total body irradiation. MDS, myelodysplastic syndromes; MPN, myeloproliferative neoplasms.
*\*The in vivo T-cell depletion category includes anti-thymocyte globulin or anti-T-lymphocyte globulin (ATG/ATLG) only, without post-transplant cyclophosphamide (PTCy).*

### Impact of the single HLA mismatches on post-HCT outcomes

Starting from this cohort, we first examined overall transplant outcomes to contextualize subsequent analyses. The median follow-up was 57.23 months (interquartile range [IQR], 35.71– 83.91 months). The 2-year OS probability was 54.3% (95% CI, 52.8–55.8), the 2-year RFS was 48.8% (95% CI, 47.3–50.3), and the 2-year NRM was 23.7% (95% CI, 22.5–25.0) (Figure S2A–B). The cumulative incidence of grade II-IV acute GvHD by day 100 was 31.6% (95% CI, 30.2–32.9), while the cumulative incidence of extensive chronic GvHD at 2 years was 36.0% (95% CI, 34.5–37.4). We then performed univariable analyses to assess the impact of individual HLA mismatches on these outcomes. Each mismatched locus was associated with a distinct effect on survival and event rates. Class I mismatches (HLA-A, -B, and -C) were associated with poorer survival, with HLA-A mismatches conferring the greatest risk for both inferior OS and RFS, whereas mismatches at HLA-DRB1 and DQB1 were associated with comparatively less detrimental outcomes (p<0.001). A similar pattern was observed for NRM, with significantly higher mortality rates in class I–mismatched transplants compared with DRB1- and DQB1-mismatched transplants, and the lowest incidence of NRM observed in DRB1 mismatches (p<0.001). In contrast, the cumulative incidence of relapse did not differ significantly across mismatch groups (p=0.36), suggesting a limited impact of HLA mismatching on relapse risk. The cumulative incidence of acute GvHD was significantly higher in case of class I mismatches, particularly at HLA-A and HLA-C, whereas DQB1 mismatches were associated with a lower incidence (p=0.01). Chronic GvHD showed a similar trend, with higher cumulative incidence in class I–mismatched transplants, although this difference did not reach statistical significance (p=0.064) (**Figure 2A**; Table S1).

**Figure 2.**
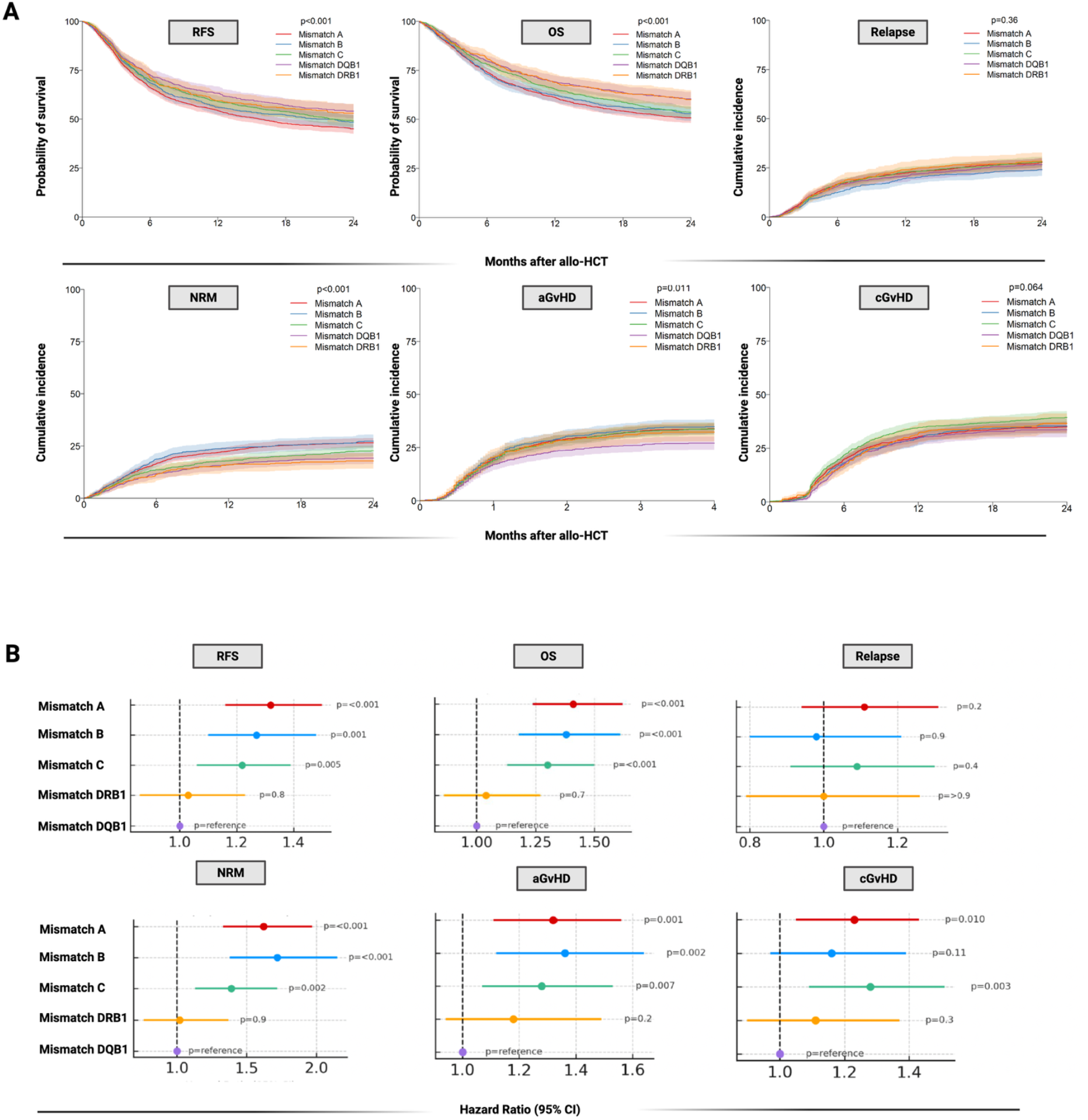
Locus-specific impact of HLA mismatches on post-transplant outcomes in 9/10 MMUD allo-HCT. **(A)** Unadjusted outcome curves stratified by the mismatched HLA locus (HLA-A, -B, -C, -DQB1, - DRB1). Relapse-free survival (RFS) and overall survival (OS) are shown as Kaplan–Meier estimates, whereas relapse, non-relapse mortality (NRM), acute GvHD grade II–IV (aGvHD), and chronic GvHD (cGvHD) are shown as cumulative incidence functions accounting for competing risks (death for relapse; relapse for NRM). Shaded bands indicate 95% confidence intervals; p-values denote overall differences between locus-specific groups (based on log-rank or Gray test). Time is displayed as months after allo-HCT. **(B)** Multivariable analyses of the association between mismatched HLA locus and each endpoint, reported as hazard ratios (HR) with 95% confidence intervals from cause-specific Cox models. HLA-DQB1 mismatch was used as the reference category. Models were adjusted for stem-cell source, patient age (per decade), donor age (per decade), Disease Risk Index, T-cell depletion, donor sex match, Karnofsky score, CMV serostatus, year of transplant, total body irradiation, conditioning intensity, and GvHD prophylaxis.

After adjustment for relevant baseline covariates (*Supplementary Methods*), locus-related differences in outcomes remained consistent with univariable analyses. Class I mismatches were independently associated with inferior survival, RFS, higher NRM and increased risk of acute and chronic GvHD, with the strongest and most consistent effects observed for HLA-A and HLA-B mismatches, and more moderate effects for HLA-C. In contrast, DQB1 and DRB1 mismatches were largely neutral across these endpoints, confirming their comparatively limited detrimental impact after multivariable adjustment (**Figure 2B, Table S2**).

### Impact of intra and interindividual HED metrics on post-HCT outcomes

Building on these observations, we next examined whether locus-specific recipient and donor HED, as well as interindividual HED at the mismatched locus, could further refine outcome prediction within each mismatch subgroup. To this end, we applied a multivariable framework incorporating clinically relevant covariates and used cause-specific Cox regression models to assess associations between HED metrics and survival outcomes or cumulative event risks. Separate models were constructed for each endpoint and each HED metric, and analyses were performed within individual mismatch strata to allow for locus-specific associations and potential cross-locus interactions (**Figure 3A,B, Tables S3**). Overall, HED-based metrics provided a finer stratification of post-transplant outcomes than dichotomous allele-level mismatch classification alone, with the most consistent and clinically relevant associations involving class II–related metrics. This was particularly evident in the DRB1-mismatched subgroup, which overall displayed comparatively favorable outcomes. Within this group, greater interindividual DRB1 HED-MM was independently associated with inferior RFS immediately after transplant (HR 1.14, 95% CI 1.02–1.28; p=0.021, for unit increase of HED-MM score), with a progressive attenuation of this association over time (time interaction HR 0.94, 95% CI 0.89–0.99; p=0.026 at 2 years) (**Figure 3B**). Recipient DRB1 HED also showed a modest but significant association with worse RFS (HR 1.04, 95% CI 1.00–1.08; p=0.042), whereas no significant DRB1-related effects were observed for OS, relapse incidence, NRM, acute or chronic GvHD. Across other mismatch strata, we observed cross-locus associations, whereby recipient class II HED influenced outcomes in class I–mismatched settings, and vice versa. First, in B-mismatched pairs, higher recipient divergence at class II loci (particularly DRB1) was consistently associated with adverse outcomes, including inferior RFS (HR 1.03, 95% CI 1.01–1.05; p=0.016), worse OS (HR 1.03, 95% CI 1.01–1.06; p=0.007) and increased relapse risk (HR 1.03, 95% CI 1.00–1.07; p=0.046). In contrast, within the same B-mismatch subgroup, higher recipient HED at DQB1 appeared protective, associating with improved RFS (HR 0.98, 95% CI 0.96–1.00; p=0.046) and OS (HR 0.98, 95% CI 0.96– 1.00; p=0.027), highlighting divergent and locus-specific associations within the class II region (**Table S3**). Within DRB1-mismatched transplants, higher recipient HED at HLA-C was associated with increased NRM early after transplant (HR 1.32, 95% CI 1.00–1.73; p=0.047), with this effect diminishing over time (time interaction HR 0.85, 95% CI 0.75–0.96; p=0.011) and higher HED-R in locus A associated with increased risk of acute GvHD (HR 1.07 95%CI 1.01-1.13, p=0.015)

**Figure 3.**
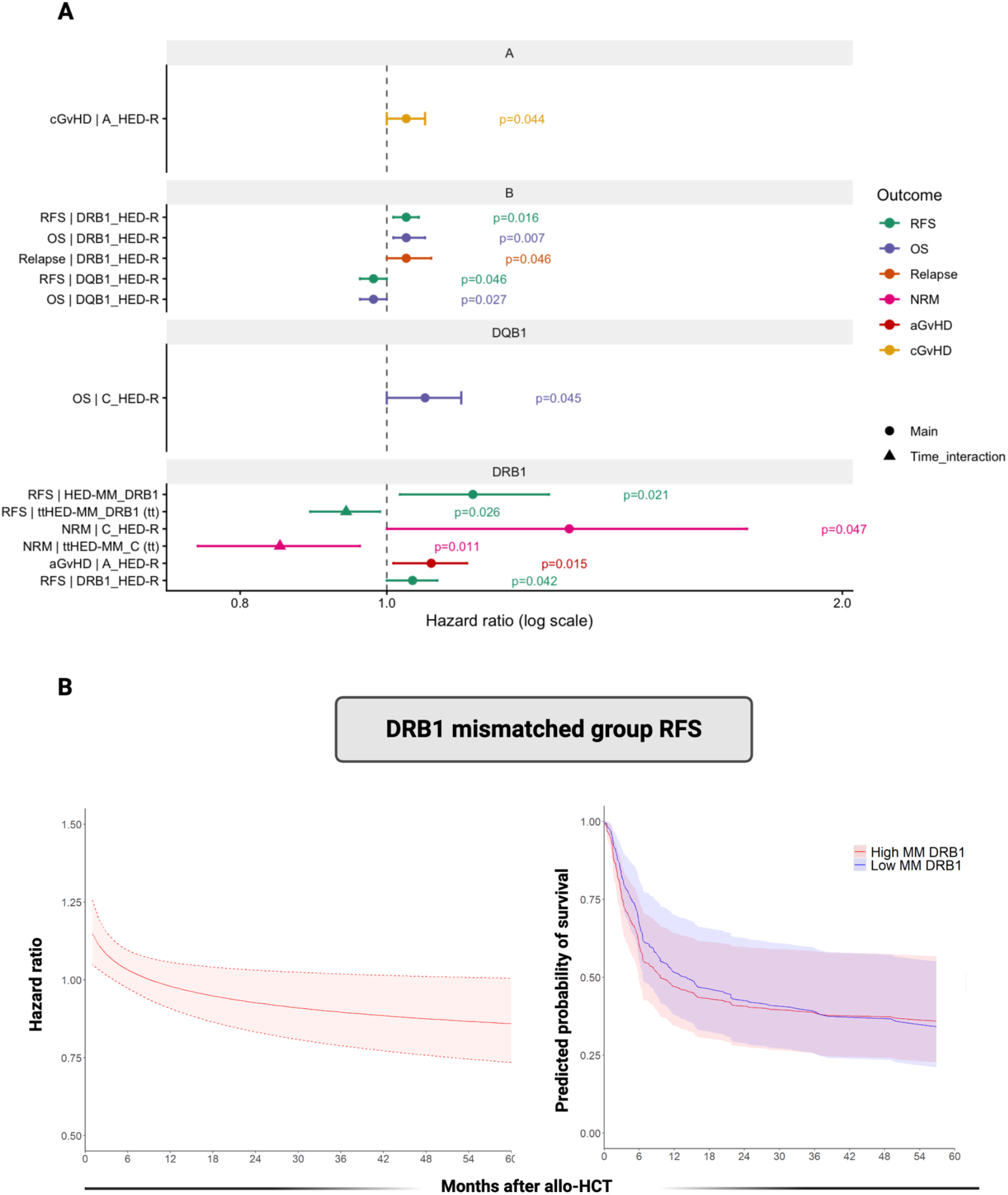
Significant associations of intra- and inter-individual HED metrics with post–allo-HCT outcomes and time-varying effect in the DRB1-mismatched subgroup. **(A)** Forest plot summarizing all statistically significant associations identified in multivariable cause-specific Cox models within each mismatch stratum (HLA-A, -B, -DQB1, and -DRB1), full results are reported in table S3. Points represent hazard ratios (HR) and horizontal bars 95% confidence intervals on a log scale; the vertical dashed line indicates HR=1. Colors denote the evaluated endpoint (RFS, OS, relapse, NRM, aGvHD, cGvHD). Circles indicate main (time-constant) effects, whereas triangles denote terms modeled with a time-interaction (time-dependent association). **(B)** Time-varying association between interindividual DRB1 HED mismatch (HED-MM_DRB1) and RFS in the DRB1-mismatched subgroup. Left: estimated time-dependent HR function with 95% confidence band, showing attenuation of the adverse effect over time. Right: model-based predicted RFS for high vs low HED-MM_DRB1 (dichotomized), with shaded 95% confidence bands. Models were adjusted for the clinical covariates described in Methods/Table S3. The model was adjusted for stem-cell source, patient and donor age, year of transplant, DRI, T-cell depletion, donor sex match, Karnofsky score, CMV status, total body irradiation, conditioning intensity, and GvHD prophylaxis. Low and High Mismatch DRB1 are defined as the 1st quartile point (Low) and 3rd quartile point (High).

Similarly, in the DQB1-mismatched subgroup, higher recipient HED at HLA-C was associated with inferior OS (HR 1.06, 95% CI 1.00–1.12; p=0.045). Within-class interactions were also observed for class I: indeed, in A-mismatched pairs, higher HED-R at HLA-A was associated with an increased risk of chronic GvHD (HR 1.03, 95% CI 1.00–1.06; p=0.044).

By contrast, no significant associations were observed within the C-mismatch subgroup across endpoints, and interindividual HED-mismatch metrics outside DRB1 were largely neutral. Together, these results indicate that HED metrics uncover clinically meaningful, locus-dependent heterogeneity within conventional mismatch categories and context-dependent cross-locus interactions.

### HLA-DPB1 focused models

We next asked whether, within the 9/10 HLA-mismatched setting, DPB1 variation refined outcome associations. Within the subset of patients with a DPB1 mismatch in addition to a single class I or class II mismatch (N = 1389, **Table S4**), neither recipient DPB1 HED (HED-R) nor interindividual DPB1 HED-mismatch (HED-MM) were associated with post-transplant outcomes. In multivariable cause-specific Cox models adjusted for clinical covariates, DPB1 HED-R showed no significant association with overall survival, relapse-free survival, relapse incidence, non-relapse mortality, acute GvHD grade II–IV, or chronic GvHD (all p > 0.40). Similarly, the magnitude of DPB1 HED-MM was not associated with any evaluated endpoint, including overall survival (HR 1.00, 95% CI 0.98–1.01; p = 0.50), relapse-free survival (HR 1.00, 95% CI 0.98–1.01; p = 0.50), relapse (HR 1.01, 95% CI 0.98– 1.02; p = 0.90), non-relapse mortality (HR 1.01, 95% CI 0.99–1.01; p = 0.40), acute GvHD II–IV (HR 1.00, 95% CI 0.98–1.01; p = 0.50), or chronic GvHD (HR 0.99, 95% CI 0.97–1.01; p = 0.40). No time-dependent associations were detected for DPB1-related metrics across outcomes.

## Discussion

Advancements in immunogenetics have highlighted the need to move beyond a binary classification of HLA mismatching towards a more functional characterization of immunogenetic disparities in allo-HCT^5^. Despite these advances, a comprehensive understanding of the functional role of each mismatched locus remains an unmet need in allo-HCT. In this study, we leveraged HED, a refined metric of HLA structural diversity, to dissect the immunogenetic nuances and clinical implications of each specific mismatch within a well-defined and homogeneous cohort of patients undergoing allo-HCT from a mismatched unrelated donor for selected hematological malignancies. We provide a unique immunogenetic benchmark to explore donor-recipient interactions, demonstrating how different HLA configurations within the mismatched loci can influence post-transplant outcomes, suggesting that interindividual HED variations at mismatched loci may contribute to immunological responses post-transplantation. Our findings indicate that the biological consequences of HLA mismatching are not uniform across class I and class II loci. Beyond the mere presence of a mismatch, qualitative differences in donor–recipient HLA divergence appear to capture meaningful variation in the allogeneic “distance” that shapes post-transplant outcomes. Taken together, these data support a shift from treating HLA disparities as binary events toward conceptualizing them as biologically graded perturbations of antigen processing and peptide presentation, with locus- and context-specific associations on alloreactivity.

Consistent with recent EBMT and CIBMTR studies^10,11^, we confirm here that not all single HLA mismatches exert equivalent clinical effects. In particular, class I mismatches, especially at HLA-A and HLA-B, were independently associated with higher risks than mismatches at DRB1 and DQB1. Importantly, this hierarchy persisted in multivariable analyses, reinforcing the robustness of locus-specific associations. The comparatively benign (less detrimental) profile of DRB1 mismatches contrasts with longstanding assumptions in the transplantation literature, which have traditionally ranked DRB1 among the most deleterious loci^8^. Our results suggest that DRB1 mismatching per se is not uniformly harmful. Rather, this locus appears to harbor substantial internal heterogeneity, which becomes apparent only when qualitative features of the mismatch are considered. This observation provides a biological rationale for the inconsistent effects of DRB1 mismatching reported across historical cohorts and underscores the limitations of allele-level mismatch labels alone^8,21^. Indeed, although DRB1 mismatches were globally associated with more favorable outcomes (among other mismatch categories), higher interindividual DRB1 HED-mismatch independently predicted inferior RFS early after transplant, with attenuation over time. This time-dependent association suggests that early post-transplant immune dynamics (when donor T-cell priming against recipient antigens is most intense) may be particularly sensitive to the degree of immunopeptidome divergence at this locus. Recipient DRB1 HED also showed a modest association with RFS, further supporting a role for class II diversity in shaping GvT effects without necessarily exacerbating GvHD.

Beyond DRB1, our analyses uncovered cross-locus effects across mismatches whereby recipient class II HED influenced outcomes in class I–mismatched settings, and vice versa. These findings argue against a strictly locus-isolated view of alloreactivity and instead support a model in which the global HLA divergence (and indirectly, immunopeptidome) landscape modulates the clinical impact of a given mismatch. One possible mechanistic explanation for the cross-locus interactions is that polymorphisms at the mismatched HLA locus generate a repertoire of donor HLA–derived peptides that can be processed and presented by shared/matched HLA molecules, thereby behaving functionally as minor histocompatibility antigens and amplifying indirect allorecognition. Along the same conceptual line, peptide-centric immunogenetic models (most notably the Predicted Indirectly ReCognizable HLA Epitopes (PIRCHE) algorithm) aim to quantify the burden of donor HLA– derived peptides predicted to be indirectly presented by recipient–donor shared HLA molecules, providing an operational estimate of indirect alloreactive load^27^. However, the clinical signal and transportability of such indices appear context-dependent, with heterogeneous applicability across transplant platforms and immune-modulation strategies^28–30^. Our study expands upon previous research by shifting from traditional allele-level mismatching to a more nuanced analysis of immunogenetic diversity. While conventional HLA matching models have primarily focused on allele-level compatibility, growing evidence indicates that HLA polymorphisms influence antigen presentation and T-cell repertoire diversity in ways that binary matching approaches fail to capture^31,32^. The integration of the HED metrics provides a novel framework for quantifying these immunogenetic differences and assessing their functional impact on transplant outcomes.

Our focused analyses of DPB1 provide additional insight. Neither DPB1 HED-R nor interindividual DPB1 divergence was associated with outcomes in our 9/10 MMUD cohort. These findings suggest that in the context of an existing class I or class II mismatch, DPB1-related divergence may exert limited incremental influence, or its effects may already be subsumed by other immunogenetic factors. Besides, we need to take into account that DPB1 locus, despite harboring a lower number of polymorphisms, is frequently mismatched, which in our cohort resulted in a divergence distribution shifted toward a reduced discriminative range, potentially limiting power to detect gradient effects. Moreover, DPB1-driven alloreactivity may be influenced by qualitative mismatch features (including permissiveness frameworks and expression-related effects)^22–26^ that may not be fully captured by a global peptide-binding divergence metric.

Several limitations merit consideration. The retrospective design and reliance on registry data preclude direct assessment of antigen specificity, T-cell clonality or other functional measurements. Our analyses span a prolonged study period and therefore may be influenced by temporal heterogeneity in transplant practices. In addition, by design, our study was restricted to transplants performed without PT-Cy. While this approach limits the generalizability of our findings to contemporary transplant platforms in which PT-Cy is routinely employed, it allowed us to isolate the net impact of individual HLA mismatches without the confounding immunomodulatory effects of PT-Cy-based T-cell repletion^33–38^. Additionally, while HED provides a structured proxy for immunopeptidome diversity, it does not capture peptide processing, expression levels, or T-cell receptor recognition. A further limitation is that HED is computed across the entire antigen-binding domain with equal weighting of all amino-acid positions; it does not discriminate residues that directly shape the peptide-binding groove (or TCR-facing surfaces) from positions with limited functional contribution, and may therefore dilute the influence of key polymorphisms that drive peptide specificity and immunogenicity. Finally, because within-locus HED is defined by pairwise divergence between the two alleles, homozygous genotypes are assigned a value of zero by construction; this collapses biologically distinct homozygous states into a single category and does not account for potential differences in functional divergence across alleles in the broader population.

From a clinical perspective, our results may have direct implications for donor selection in MMUD transplantation in non-PT-Cy platforms, which are still routinely used in many centers especially in Europe. First, HED-based metrics could be integrated into unrelated donor selection algorithms to refine risk stratification among otherwise equivalent 9/10 MMUD options, particularly within DRB1-mismatched settings. Rather than selecting donors based solely on the presence of a mismatch, divergence-based information may help identify permissive versus high-risk mismatches and support more individualized donor choice. In addition, HED metrics could inform post-transplant risk assessment, identifying patients who may benefit from closer monitoring or tailored immunosuppressive or immunomodulatory strategies (including donor lymphocyte infusion^39^) in the early post-transplant period.

In summary, this study demonstrates that HED-based metrics complement conventional HLA mismatch classification by capturing biologically meaningful heterogeneity in donor-recipient immunopeptidome divergence. By revealing locus-dependent and cross-locus effects, particularly within class II mismatches, our findings support a more refined approach to donor selection in MMUD allo-HCT and lay the groundwork for future precision immunogenetic strategies, including exploration of PT-Cy effects.

## Data Availability

All the data that support the findings of this study are available within the Article and Supplementary Files. The final dataset can be made available to all interested researchers upon request to CTIWP chair and data management team.

## Authorship contributions

SP, TL, CG, KF and AR conceptualized the study, designed the metrics and interpreted the data analysis. SP provided the study synopsis, wrote the manuscript and designed the study and the statistical plan. TL performed HED-related metrics’ computation. JEM contributed to metric design, to the discussion of analytical plan and performed biostatistical analyses. FA, RZ, VP, PD, WB, IH, DM, AR, HS, JP, DR, TGD, FK, ME, SM, ME were the PIs of the centers mainly involved in patient recruitment and performed data-collection at the center level through EBMT registry. MA, PC, PM, CC, JK, LCDW participated to study conception and to the interpretation of the data analysis and provided important intellectual inputs. JDH performed data management and coordinated the study at the EBMT level. TL, KF and AR supervised the study, interpreted the data analysis, gave important intellectual inputs and edited the manuscript. All authors reviewed and approved the final version of this manuscript.

## Acknowledgements

This work was supported by EBMT. We acknowledge all participating centers. TLL was supported by the Deutsche Forschungsgemeinschaft (DFG, German Research Foundation) – 437857095. SP received funding from Fondation ARC pour la Recherche sur le Cancer, Force Hemato. KF was supported by the Deutsche José Carreras Leukemie Stiftung (DJCLS 17 R/2023).

