## Supplementary appendix for "Interindividual HLA Evolutionary Divergence in Single HLA-Mismatched Unrelated Donor Hematopoietic Cell Transplantation for Malignant Hematological Disorders: A Report on Behalf of the Cellular Therapy and Immunobiology Working Party of the EBMT"

*Co-last authors

#### List of EBMT centers participating in the study

Thomas Schroeder, University Hospital | Essen, Essen, Germany;

Francis Ayuk, University Hospital Eppendorf, Hamburg, Germany;

Robert Zeiser, University of Freiburg, Freiburg, Germany;

Victoria Potter, Kings College Hospital London, London, United Kingdom;

Peter Dreger, University of Heidelberg, Heidelberg, Germany;

Wolfgang Bethge, Universitaet Tuebingen, Tuebingen, Germany;

Inken Hilgendorf, Universitaetsklinikum Jena, Jena, Germany;

Regis Peffault de Latour, Saint-Louis Hospital, BMT Unit, Paris, France;

Alessandro Rambaldi, ASST Papa Giovanni XXIII, Bergamo, Italy;

Henrik Sengeloev, Rigshospitalet, Copenhagen, Denmark;

Jakob Passweg, University Hospital | Basel, Basel, Switzerland;

Deborah Richardson, Southampton General Hospital, Southampton, United Kingdom;

Tobias Gedde-Dahl, Oslo University Hospital, Rikshospitalet, Oslo, Norway;

Francesca Kinsella, Birmingham Centre for Cellular Therapy and Transplant (BCCTT), Birmingham, United Kingdom;

Matthias Edinger, University Regensburg, Regensburg, Germany;

Stephan Mielke, Karolinska University Hospital, Stockholm, Sweden;

Matthias Eder, Hannover Medical School, Hannover, Germany;

Pavel Jindra, Charles University Hospital, Pilzen, Czech Republic;

Charles Crawley, Addenbrookes Hospital Cambridge, Cambridge, United Kingdom;

Helene Labussiere-Wallet, Centre Hospitalier Lyon Sud, Pierre BΘnite, France;

Jan Vydra, Institute of Hematology and Blood Transfusion, Prague, Czechia;

Ibrahim Yakoub-Agha, CHU de Lille, Lille, France;

Jennifer Byrne, Nottingham City Hospital, Nottingham, United;

Johan Maertens, University Hospital Gasthuisberg, Leuven, Belgium;

Alessandro Busca, S.S.C.V.D Trapianto di Cellule Staminali, Torino, Italy;

Urpu Salmenniemi, HUCH Comprehensive Cancer Center, Helsinki, Finland;

Pavel Zak, Charles University Hospital, Hradec, Czech Republic;

Katherine Clesham, University College London Hospital, London, United Kingdom;

Matteo Parma, Ospedale San Gerardo, Monza, Italy;

Ladislav Sopko, University Hospital, Bratislava, Slovak Republic;

Martin Bornhauser, Universitaetsklinikum Dresden, Dresden, Germany;

Andrew Clark, Glasgow Royal Infirmary, Glasgow, United Kingdom;

Francesca Bonifazi, IRCCS Azienda Ospedaliero-Universitaria di Bologna, Bologna, Italy;

Martin Kaufmann, Robert_Bosch_Krankenhaus, Stuttgart, Germany;

Carlos Manuel Panizo Santos, Hospital Universitario Donostia, S Sebastian, Spain;

Xavier Poire, Cliniques Universitaires St. Luc, Brussels, Belgium;

Marco Zecca, San Matteo Pavia Transplant Programme, Pavia, Italy;

Lorenz Thurner, University of Saarland, Homburg, Germany;

Kristina Carlson, University Hospital | Uppsala, Uppsala, Sweden;

Didier Blaise, Programme de Transplantation&Therapie Cellulaire, Marseille, France;

Keith Wilson, Cardiff University Hospital of Wales & Swansea, Cardiff, United Kingdom;

Alexander Kulagin, RM Gorbacheva Research Institute, Pavlov University, St. Petersburg, Russian Federation (the);

Dominik Schneidawind, University Hospital, Zⁿrich, Switzerland;

Friedrich Stolzel, University Medical Center Schleswig-Holstein, Campus Kiel, Kiel, Germany;

Eleni Tholouli, Manchester Royal Infirmary, Manchester, England;

Edouard Forcade, CHU Bordeaux, Hopital Haut-Leveque, Pessac, France;

Anne Huynh, CHU - Institut Universitaire du Cancer Toulouse, Toulouse, France;

Cristina Castilla-Llorente, Gustave Roussy Cancer Campus, Villejuif, France;

Marie Therese Rubio, CHRU NANCY, Vandoeuvre les Nancy, France;

Jrgen Kuball, University Medical Centre Utrecht, Utrecht, Netherlands (the);

Mieke Roeven, Nijmegen Medical Centre, Nijmegen, Netherlands;

Domenico Russo, USD Trapianti di Midollo, Adulti, Brescia, Italy;

Alexandros Spyridonidis, Research Committee - University of Patras, Patras, Greece;

Polina Stepensky, Hadassah University Hospital, Jerusalem, Israel;

Gwendolyn Van Gorkom, University Hospital Maastricht, MAASTRICHT, Netherlands (the);

Claude Eric Bulabois, CHU Grenoble Alpes - Universite Grenoble Alpes, Grenoble, France;

Patrice Chevallier, CHU Nantes, Nantes, France;

Gandhi Damaj, CHU CAEN, Caen, France;

Georg-Nikolaus Franke, Medical Clinic and Policinic 1, Leipzig, Germany;

Kazimierz Halaburda, Institute of Hematology and Transfusion Medicine, Warsaw, Poland;

Stig Lenhoff, Skanes University Hospital, Lund, Sweden;

Emma Nicholson, Royal Marsden Hospital, London, United Kingdom;

Ludek Raida, Olomouc University Hospital, Olomouc, Czech Republic;

Annoek Broers, Erasmus MC Cancer Institute, Rotterdam, Netherlands;

Sebastien Maury, Hopital Henri Mondor, Creteil, France;

Jose Antonio Perez-Simon, Hospital Universitario Virgen del Rocio, Sevilla, Spain;

Peter Remenyi, Del-pesti Centrumkorhaz, Budapest, Hungary;

Johanna Tischer, Klinikum Grosshadern, Munich, Germany;

Francesco Zallio, H SS. Antonio e Biagio, Alessandria, Italy;

Thomas Cluzeau, CHU Nice - Hopital de l'ARCHET I, Nice, France;

Carlos Pinho Vaz, Inst. Portugues de Oncologia do Porto, Porto, Portugal;

Omur Gokmen Sevindik, Demiroglu Bilim University Istanbul Florence Nightingale Hospital, Istanbul, Turkey;

John Snowden, Sheffield Royal Hallamshire, Sheffield, United Kingdom;

Frederic Baron, University of Liege, Liege, Belgium;

Irene Cavattoni, Hospital San Maurizio, Bolzano, Italy;

Xavier Leleu, Hopital La Miletrie, Poitiers, France;

Arancha Bermudez Rodriguez, Hospital U. Marques de Valdecilla, Santander, Spain;

Werner Rabitsch, Medizinische Universitaet Wien, Vienna, Austria;

Raffaella Cerretti, Tor Vergata University of Rome, Rome, Italy;

Christof Scheid, University of Cologne, Cologne, Germany;

Mareike Verbeek, TUM Universitatsklinikum, Klinikum rechts der Isar, Munich, Germany;

Peter A. von dem Borne, Leiden University Hospital, Leiden, Netherlands;

Ahmet Elmaagacli, Asklepios Klinik St. Georg, Hamburg, Germany;

John. G. Gribben, St. Bartholomew`s Hospital London, London, United Kingdom;

Jan-Erik Johansson, Sahlgrenska University Hospital, Goeteborg, Sweden;

Cristina Tecchio, Azienda Ospedaliera Universitaria Integrata Verona, Verona, Italy;

Fabio Ciceri, IRCCS San Raffaele Hospital Scientific Institute, Milano, Italy;

Zubeyde Nur Ozkurt, Gazi University Faculty of Medicine, Ankara, Turkey;

Elisa Sala, Klinik fuer Innere Medzin III, Ulm, Germany;

Christoph Schmid, Klinikum Augsburg, Augsburg, Germany;

Anna Torrent Catarineu, ICO-Hospital Universitari Germans Trias i Pujol, Badalona, Spain;

Tsila Zuckerman, Rambam Medical Center, Haifa, Israel;

Anna Paola Iori, Univ. La Sapienza, Rome, Italy;

Philippe Lewalle, Institut Jules Bordet, Brussels, Belgium;

Ron Ram, Tel Aviv Sourasky Medical Center, Tel Aviv, Israel;

Eva Wagner-Drouet, University Medical Center Mainz, Mainz, Germany;

Jacques-Olivier Bay, CHU ESTAING, Clermont, France;

Carlo Borghero, S. Bortolo Hospital, Vicenza, Italy;

Renato Fanin, Azienda Sanitaria Universitaria Friuli Centrale, Udine, Italy;

Lucia Lopez Corral, Hospital Clinico, Salamanca, Spain;

Stephanie Nguyen Quoc, Universite Paris IV, Hopital la Pitie-Salpetriere, Paris, France;

Franca Fagioli, Ospedale Infantile Regina Margherita, Torino, Italy;

Catherine Flynn, Hope Directorate - St. James's Hospital, Dublin, Ireland;

Maija Itala-Remes, Turku University Hospital, Turku, Finland;

Gitte Olesen, Aarhus University Hospital | Denmark, Aarhus N, Denmark;

Attilio Olivieri, Azienda Ospedali Riuniti di Ancona, Ancona, Italy;

Anne Sirvent, CHU Lapeyronie, Montpellier, France;

Carin Hazenberg, University Medical Center Groningen (UMCG), Groningen, Netherlands (the);

Cecilia Isaksson, Umea University Hospital, Umeσ, Sweden;

Jiri Mayer, University Hospital Brno, Brno, Czech Republic;

Erfan Nur, Amsterdam UMC, Amsterdam, Netherlands;

Eric Deconinck, Hopital Jean Minjoz, Besancon, France;

Olivier Hermine, Hopital Necker Adults, Paris, France;

Patrick Medd, Derriford Hospital Plymouth, Plymouth, United Kingdom;

Yves Chalandon, Departement d'Oncologie, Service d'Hematologie, Geneva, Switzerland;

Tom Lodewyck, A.Z. Sint-Jan, Brugge, Belgium;

Jan Zaucha, University Clinical Centre in Gdansk, Gdansk, Poland;

Giovanni Grillo, ASST GRANDE OSPEDALE METROPOLITANO NIGUARDA, Milano, Italy;

Andy Peniket, Oxford Radcliffe, Oxford, United Kingdom;

Montserrat Rovira, Hospital Clinic, Barcelona, Spain;

Kerstin Schafer-Eckart, Klinikum Nuernberg, Nuernberg, Germany;

Nathalie Contentin, Centre Henri Becquerel, Rouen, France;

Marieke Drholt, Evangelisches Krankenhaus Essen-Werden gGmbH, Essen, Germany;

Hermann Einsele, Universitaetsklinikum Wuerzburg, Wuerzburg, Germany;

Piero Galieni, Mazzoni Hospital, Ascoli Piceno, Italy;

Matthias Stelljes, University of Muenster, Muenster, Germany;

Dominik Wolf, University Hospital Innsbruck, Innsbruck, Austria;

Ipek Yonal-Hindilerden, Istanbul Tip Fakultesi, Istanbul, Turkey;

Adrian Bloor, Christie Hospital Manchester, Manchester, United Kingdom;

Mathilde Hunault-Berger, CHRU, Angers, France;

Fernando Leal da Costa, Inst. Portugues Oncologia, Lisboa, Portugal;

Alexander Murray Martin, Leicester Royal Infirmary, Leicester, United Kingdom;

Lucia Prezioso, Univ. of Parma, Parma, Italy;

Shankara Paneesha, Birmingham Centre for Cellular Therapy and Transplant (BCCTT), Birmingham, United Kingdom;

Jaroslaw Dybko, Department of Hematology and Transplantology of Lower Silesian Center of Oncology, Wroclaw, Poland;

Alain Gadisseur, Antwerp University Hospital (UZA), Antwerp, Belgium;

Edgar Jost, University Hospital Aachen, Aachen, Germany;

Tessa Kerre, Ghent University Hospital, Gent, Belgium;

Nicola Mordini, Az. Ospedaliera S. Croce e Carle, Cuneo, Italy;

Depei Wu, First Affiliated Hospital of Soochow University, Suzhou, China;

Grzegorz Basak, Central Clinical Hospital, Warsaw, Poland;

Edoardo Benedetti, Azienda Ospedaliero Universitaria Pisana, Pisa, Italy;

Anna Bergendahl Sandstedt, University Hospital | Linkoeping, Linkoeping, Sweden;

Angelo Michele Carella, IRCCS, Casa Sollievo della Sofferenza, San Giovanni, Italy;

Paola Carluccio, U.O. Ematologia con Trapianto, Bari, Italy;

Luca Castagna IT, U.O.D Trapianti di midollo osseo, Palermo, Italy;

Angela Cuoghi, Azienda Ospedaliero Universitaria di Modena Policlinico, Modena, Italy;

Gaelle Guillerm, C.H.R.U de Brest, Brest, France;

Ain Kaare, Tartu University Hospital, Tartu, Estonia;

Giorgio La Nasa, Centro Trapianti Unico Di CSE Adulti e Pediatrico A. O Brotzu, Cagliari, Italy;

Bruno Lioure, ICANS - Institut de cancerologie Strasbourg Europe, Strasbourg, France;

Elena Parovichnikova, National Research Center for Hematology, Moscow, Russian Federation;

Ioanna Sakellari, George Papanicolaou General Hospital, Thessaloniki, Greece;

Teresa Zudaire, Hospital Universitario de Navarra, Pamplona, Spain;

Ann De Becker, Universitair Ziekenhuis Brussel, Brussels, Belgium;

Dries Deeren, AZ Delta, Roeselare, Belgium;

Alessandro Maggi, Ospedale Nord, Taranto, Italy;

Stella Santarone, Ospedale Civile, Pescara, Italy;

Matjaz Sever, University Med. Center, Ljubljana, Slovenia;

Alina Tanase, Fundeni Clinical Institute Adults, Bucharest, Romania;

Pascal Turlure, CHRU Limoges, Limoges, France;

Friederike Wortmann, University Medical Center Schleswig-Holstein, Luebeck, Germany;

Sebastian Giebel, Department of Bone Marrow Transplantation and Oncohematology, Gliwice, Poland;

William Krueger, Klinik fuer Innere Medizin C, Greifswald, Germany;

Jean-Valere Malfuson, Hopital D'Instruction des Armees (HIA) PERCY, Clamart, France;

Carlos Solano Vercet, Hospital Clinico de Valencia, Valencia, Spain;

Adrian Alegre Amor, Hospital de la Princesa, Madrid, Spain;

Emanuele Angelucci, IRCCS Ospedale Policlinico San Martino, Genova, Italy;

Matthew Collin, RVI Newcastle, Newcastle, United Kingdom;

Virginie Gandemer, Centre Hospitalier Universitaire de Rennes, Rennes, France;

Aloysius Ho, Singapore General Hospital, Singapore, Singapore;

Tobias Holderried, Universitaet Bonn, Bonn, Germany;

Salvatore Leotta, Ospedale Policlinico, Catania, Italy;

Francesco Onida, Fondazione IRCCS - Ca Granda, Milano, Italy;

Gerald Wulf, Universitaetsklinikum Goettingen, Goettingen, Germany;

Moshe Yeshurun, Beilinson Hospital, Petach-Tikva, Israel;

Carmen Albo Lopez, Hospital Alvaro Cunqueiro - Complejo Hospitalario Universitario de Vigo, Vigo, Spain;

Caroline Besley, University Hospitals Bristol and Weston NHSFT, Bristol, United Kingdom;

Jennifer Clay, St James University Hospital Leeds, Leeds, United Kingdom;

Laimonas Griskevicius, Vilnius University Hospital Santaros Klinikos, Vilnius, Lithuania;

Thomas Heinicke, Universitaetsklinium Magdeburg, Magdeburg, Germany;

Belen Sevillano Zamarreno, St. George`s Hospital London, London, United Kingdom;

David Valcßrcel, Hospital Vall d`Hebron, Barcelona, Spain;

Daniele Vallisa, Hospital Guglielmo da Saliceto, Piacenza, Italy;

Tomasz Wrobel, Uniwersytecki Szpital Kliniczny, Wroclaw, Poland;

Ka Lung Wu, ZNA, Antwerp, Belgium;

Lisa Pospiech, Klinikum Oldenburg, Oldenburg, Germany;

Armin Gerbitz, University Hospital Erlangen, Erlangen, Germany;

Concepcion Herrera Arroyo, Hosp. Reina Sofia, C≤rdoba, Spain;

Mohamad Mohty, Hopital Saint Antoine, Paris, France;

Maurizio Musso, Ospedale La Maddalena - Dpt. Oncologico, Palermo, Italy;

Yana Novis, Hospital Sirio-Libanes, Sao Paulo, Brazil;

Chiara Nozzoli, Azienda Ospedaliera Universitaria Careggi, Firenze, Italy;

Mark Ringhoffer, Klinikum Karlsruhe gGmbH, Karlsruhe, Germany;

Cristina Skert, Ospedale Dell'Angelo, Venezia, Italy;

Jan Styczynski, University Hospital, Collegium Medicum UMK, Bydgoszcz, Poland;

Monica Tozzi, U.O.S.A Centro Trapianti e Terapia Cellulare, Siena, Italy;

Paolo Corradini, University of Milano, Milano, Italy;

Hildegard Greinix, LKH - University Hospital Graz, Graz, Austria;

Grzegorz Helbig, Silesian Medical Academy, Katowice, Poland;

Franco Locatelli, IRRCS Ospedale Pediatrico Bambino Gesu, Rome, Italy;

Andres Sanchez Salinas, Hospital Universitario Virgen de la Arrixaca, Murcia, Spain;

Ali Ugur Ural, Ankara Bayindir Hospital, Ankara, Turkey;

Radovan Vrhovac, University Hospital Center Rebro, Zagreb, Croatia;

Olga Aleinikova, BelaRussian Federationn Research Center for Pediatric Oncology, Hematology and Immunology, Minsk, Belarus;

Jane Apperley, Imperial College Hammersmith London, London, United Kingdom;

Igor Wolfgang Blau, Medizinische Klinik m. S. Hamatologie , Onkologie und Tumorimmunologie, Berlin, Germany;

Katarzyna Drabko, Children`s University Hospital, Lublin, Poland;

Soledad Gonzalez Muniz, Hospital Universitario Central de Asturias, Oviedo, Spain;

Manuel Jurado Chacon, Hospital Univ. Virgen de las Nieves, Granada, Spain;

Guido Kobbe, Heinrich Heine Universitaet, Duesseldorf, Germany;

Angela Krackhardt, St. Franziskus Hospital, Flensburg, Germany;

Mi Kwon, Hospital Gregorio Maranon, Madrid, Spain;

Massimo Martino, Grande Ospedale Metropolitano Bianchi Melacrino Morelli - Centro Unico Trapianti A. Neri, Reggio Calabria, Italy;

Rodrigo Martino Bufarull, Hospital Santa Creu i Sant Pau, Barcelona, Spain;

Andrew McDonald, ALBERTS CELLULAR THERAPY, Pretoria, South Africa;

Anna Mele, Hospital C. Panico, Tricase, Italy;

Alberto Mussetti, Institut Catala d'Oncologia Hospital Duran i Reynals, Barcelona, Spain;

Michael Potter, The London Clinic Harley Street, London, United Kingdom;

Paul-Gerhardt Schlegel, University Children`s Hospital, Wuerzburg, Germany;

Simona Sica, Universita Cattolica S. Cuore, Rome, Italy;

Juan Pio Torres Carrete, Complejo Hospitalario de A Coruna, La Coru±a, Spain;

Giuseppe Visani, AORMN Hospital, Pesaro, Italy;

Serap Aksoylar, Ege University Pediatric BMT Centre, Izmir, Turkey;

Ioannis Baltadakis, Evangelismos Hospital, Athens, Greece;

Ali Bazarbachi, American University of Beirut Medical Center, Beirut, Lebanon;

Stefania Bramanti, Istituto Clinico Humanitas, Milano, Italy;

Gesine Bug, University Hospital Frankfurt - Goethe University, Frankfurt Main, Germany;

Andreas Burchert, Philipps Universitaet Marburg, Marburg, Germany;

Birgit Burkhardt, Universitaetsklinikum Muenster, Muenster, Germany;

Amandine Charbonnier, University of Amiens: CHU Amiens, Amiens, France;

Jerome Cornillon, Institut de Cancerologie Lucien Neuwirth, Saint Etienne, France;

Enrico Derenzini, European Institute of Oncology, Milano, Italy;

Burak Deveci, Medstar Antalya Hospital, Antalya, Turkey;

Ronit Elhasid, Tel-Aviv Sourasky Medical Center, Tel Aviv, Israel;

Maura Faraci, Institute G. Gaslini, Genova, Italy;

Marta Sonia Gonzalez Perez, Hospital Clinico Universitario, Santiago, Spain;

Jolanta Gozdzik, University Children`s Hospital in Krakow, Krakow, Poland;

Marφa Inmaculada Heras, Hospital Morales Meseguer, Murcia, Spain;

Bernd Hertenstein, Klinikum Bremen-Mitte, Bremen, Germany;

Leylagul Kaynar, Istanbul Medipol University | 445, Istanbul, Turkey;

Stefan Klein, Universitaetsmedizin Mannheim, Mannheim, Germany;

Alphan Kupesiz, Akdeniz University Medical School, Antalya, Turkey;

Javier Lopez Jimenez, Hospital Ramon y Cajal, Madrid, Spain;

Baris Malbora, Yeniyuzyil University, Gaziosmanpasa Hospital, Istanbul, Turkey;

Roland Meisel, Universitaetsklinikum, Duesseldorf, Germany;

Arnon Nagler, Chaim Sheba Medical Center, Tel-Hashomer, Israel;

Elettra Ortu la Barbera, Ospedale Santa Maria Goretti, Latina, Italy;

Giorgia Battipaglia, University of Napoli, Napoli, Italy;

Herbert Pichler, St. Anna Kinderspital, Vienna, Austria;

Arcangelo Prete, Azienda Ospedaliero Universitaria, Bologna, Italy;

Cecile Renard, Institut d`Hematologie et d`Oncologie Pediatrique, Lyon, France;

Muhammad Saif, Clatterbridge Cancer Centre Liverpool, Liverpool, United Kingdom;

Jaime Sanz Caballer, University Hospital La Fe, Valencia, Spain;

Roland Schroers, ZSIS Universitaetsklinikum Knappschaftskrankenhaus Bochum GmbH, Bochum, Germany;

Peter Svec, Pediatric University Teaching Hospital, Bratislava, Slovak Republic;

Milena Todorovic, University Clinical Center, Belgrade, Serbia;

Umit Barbaros Ure, Koc University Hospital, Istanbul, Turkey;

Table of content

#### Supplementary methods

##### Study design

This retrospective study, using multi-centre EBMT registry data on HSCT patients, investigates the role of recipient, donor, and donor-recipient inter-individual HED as immunogenetic determinants of post-transplant outcomes in the setting of MMUD allo-HCT for selected hematologic malignancies.

Eligible patients included adults diagnosed with acute myeloid leukemia (AML), acute lymphoblastic leukemia (ALL), myelodysplastic syndromes (MDS), or myeloproliferative neoplasms (MPN) who underwent their first allo-HCT from a 9/10 MMUD between January 2010 and July 2019 at EBMT-affiliated centers receiving both myeloablative (MAC) or reduced conditioning regimens (RIC). To ensure a homogeneous study population and accurately assess the immunogenetic effect of each mismatch, patients who received post-transplant cyclophosphamide (PT-Cy) as part of GvHD prophylaxis were not considered. Transplants were performed using either bone marrow (BM) or peripheral blood (PB) as the graft source. Only patients with 4-digit (2-field) HLA typing available for loci A, B, C, DRB1, and DQB1 were included. A subset of patients with DPB1 mismatches (additional to other class I or class II mismatches) were analyzed separately to explore the impact of HED in this locus on post-transplant outcomes.

Patients were excluded if they were pediatric (<18 years old), had received a second allo-HCT, underwent transplantation for disease indications not listed above or received umbilical cord blood (UCB) or mixed graft sources. This study was conducted in accordance with the Revised Helsinki Declaration, and all patients included provided informed consent for participation in observational studies according to EBMT policies.

##### HED metrics and Computation

HED metrics were computed at the University of Hamburg using a previously validated algorithm, which employs Grantham distance to measure differences between reciprocal amino acid residues at the antigen-binding site of a given allele pair^13^ (**Figure 1A**).

For the purposes of this study, we evaluated three distinct HED metrics (**Figure 1B**):

- Individual HED scores (computed for all HLA loci)

###### 1) Individual (intra-individual) HED

Computed per locus as the divergence between the two alleles carried by an individual:

- **Recipient HED (HED-R)**: $\mathrm{HED}(R_{1},R_{2})$
- **Donor HED (HED-D)**: $\mathrm{HED}(D_{1},D_{2})$

Where R1 and R2 are the two alleles carried by Recipient and D1 and D2 are the two alleles carried by the Donor. HED-D was computed for completeness and to support normalization steps in inter-individual metrics (below).

###### 2) Inter-individual HED at the mismatched locus (HED-MM)

For each donor–recipient pair, the **mismatched locus** contains one shared and one non-shared allele (single mismatch) on the donor and the recipient side. We defined:

$$\mathrm{HED}_{\text{between}}=\sum_{i=1}^{2} \sum_{j=1}^{2} \mathrm{HED}_{ij}$$

where $\mathrm{HED}_{ij}$ is the HED score between donor allele *i* and recipient allele *j*. This is the HED score calculated for all four allele combinations between recipient and donor (at the mismatch locus) (**Figure 1A**).

To account for the absolute “baseline” divergence of donor and recipient at that locus, $\mathrm{HED}_{\text{between}}$ was normalized by the individual HED values (recipient and donor) and then linearly rescaled for interpretability:

$$\mathrm{HED}\text{-}\mathrm{MM}=K\times\frac{\mathrm{HED}_{\text{between}}-(HED-R+HED-D)}{\mathrm{HED}_{\text{between}}+(HED-R+HED-D)}$$

where $K=30$ is a constant chosen a priori to map the observed range to an approximate **0–10 scale** (0 indicating no mismatch and higher values indicating increasing donor–recipient divergence at the mismatched locus). This normalization reflects that the same categorical mismatch can carry different functional meaning depending on the donor/recipient locus-specific diversity.

In terms of biological significance, individual HED scores (HED-D and HED-R) provide insights into the independent contributions of recipient or donor HED. In contrast, inter-individual HED scores (HED-MM) account for divergence at the mismatched locus between the patient’s and the donor’s HLA alleles. HED-MM quantifies the divergence at the mismatched locus and is intended to assess the difference in the composition of the immune peptidomes between donor and recipient, serving as a metric to distinguish permissive (lower score, more similar) from non-permissive (higher score, more dissimilar) mismatches. The metric is normalized to donor and recipient locus-specific HED to account for baseline allelic divergence, such that the same inter-individual divergence ($\mathrm{HED}_{between}$) can have different functional implications depending on the absolute diversity of the donor–recipient pair at that locus.

##### Statistical framework

Descriptive analyses of the study population were performed to summarize baseline characteristics.

Kaplan-Meier and cumulative incidence curves were constructed for respectively survival outcomes and outcomes with competing risks, with respectively a log-rank test and Gray’s test to evaluate significant group differences in survival and cumulative incidence curves. To assess the impact of HED metrics (HED-R and HED-MM) on post-transplant clinical outcomes, multivariable (cause-specific) Cox regression models were applied, incorporating relevant clinical covariates. Each HED metric was included in the model as continuous parameter.

Each model was applied to the following key post-transplant endpoints:

- **Overall Survival (OS):** Defined as the time from transplantation to death from any cause.
- **Relapse-Free Survival (RFS):** Defined as the time from transplantation to relapse with death from other causes considered as competing event.
- **Non-Relapse Mortality (NRM):** Defined as time to death without relapse.
- **acute GvHD (II-IV):** Defined as time to developing a grade II or higher (according to Seattle criteria)^21^ acute GvHD after allo-HCT, considering death before aGvHD as a competing risk.
- **Chronic GvHD:** Defined as time developing chronic GvHD after allo-HCT, considering death (before cGvHD) as a competing risk.
- **Relapse:** Defined as the time to experiencing a hematological relapse or progression after allo-HCT, while accounting for death (prior to relapse) and graft failure as competing risks.

Clinically relevant baseline variables included: diagnosis, adjusted disease risk index (DRI)^22^, year of transplant, patient and donor age, patient and donor CMV serostatus, ABO patient, donor sex (mis)match, Karnofsky score, GvHD prophylaxis regimen, conditioning regimen intensity (reduced vs. standard), stem cell source, use of TBI, T-cell depletion. For the adjusted DRI categories as few patients have a low or very high score, we distinguished between intermediate-low and high-very high.

For each outcome and in each mismatched subgroup, we built separate models for these immunogenetic parameters and checked for proportional hazards assumption (PHA) by examining the Schoenfeld residuals. In case non-proportionality was detected we allowed the HEDmismatch, or HED scores to vary over time (with an interaction with a log(t+1) function). P-values <0.05 were considered significant.

#### Supplementary figures

##### Figure S1

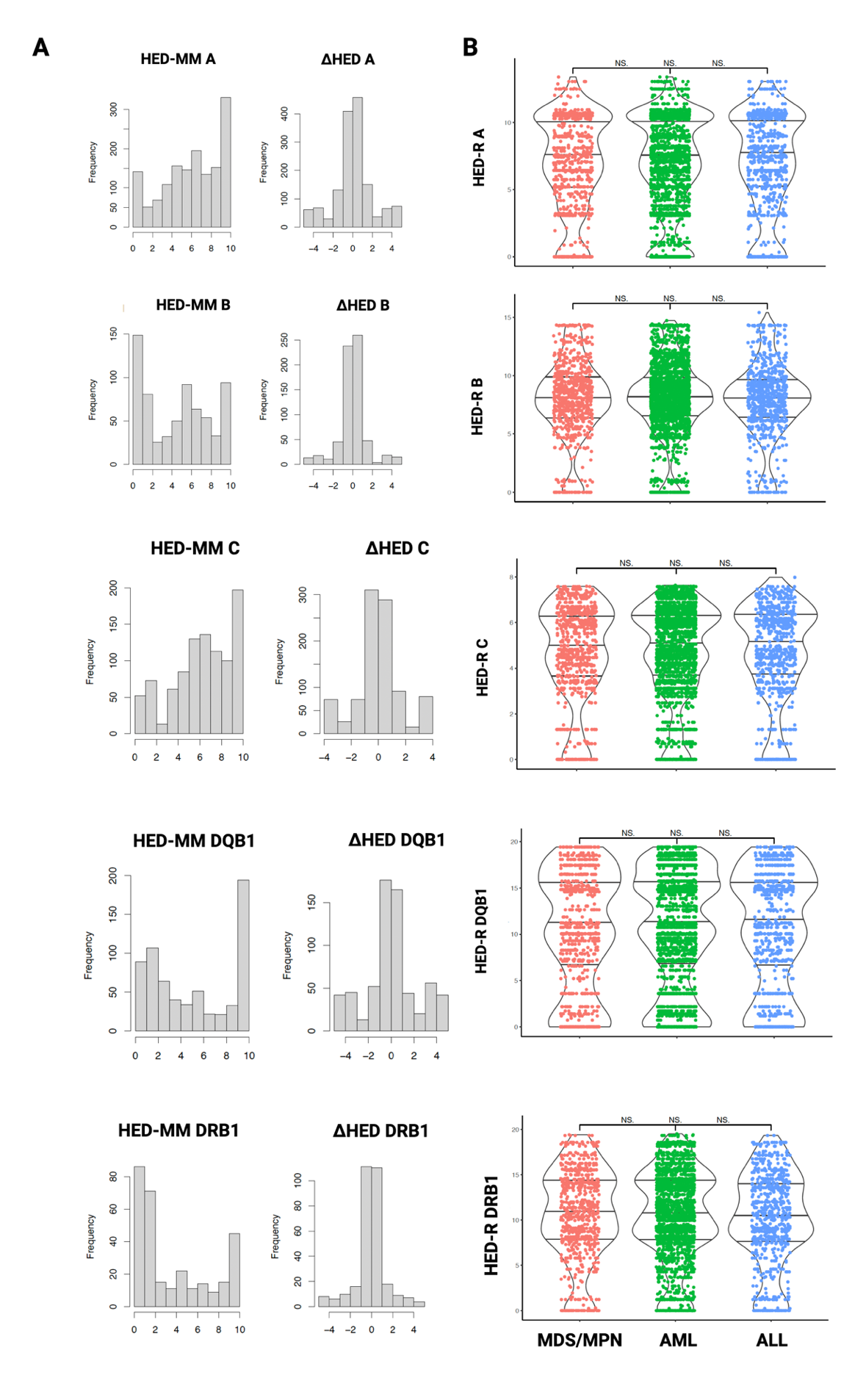

**Figure S1:** **Distribution of HED mismatch (HED-MM) and ΔHED across mismatched loci and recipient HED stratified by disease group.**

**(A)** Histograms depicting the distribution of HED-MM (left column) and ΔHED (right column) for each mismatched locus (HLA-A, -B, -C, -DQB1, -DRB1). HED-MM quantifies the immunopeptidome divergence at the mismatched locus, whereas ΔHED captures the directional difference between donor and recipient HED. **(B)** Violin plots illustrating the distribution of recipient HED (HED-R) across different hematologic malignancies: myelodysplastic syndromes/myeloproliferative neoplasms (MDS/MPN, red), acute myeloid leukemia (AML, green), and acute lymphoblastic leukemia (ALL, blue).

Figure S2
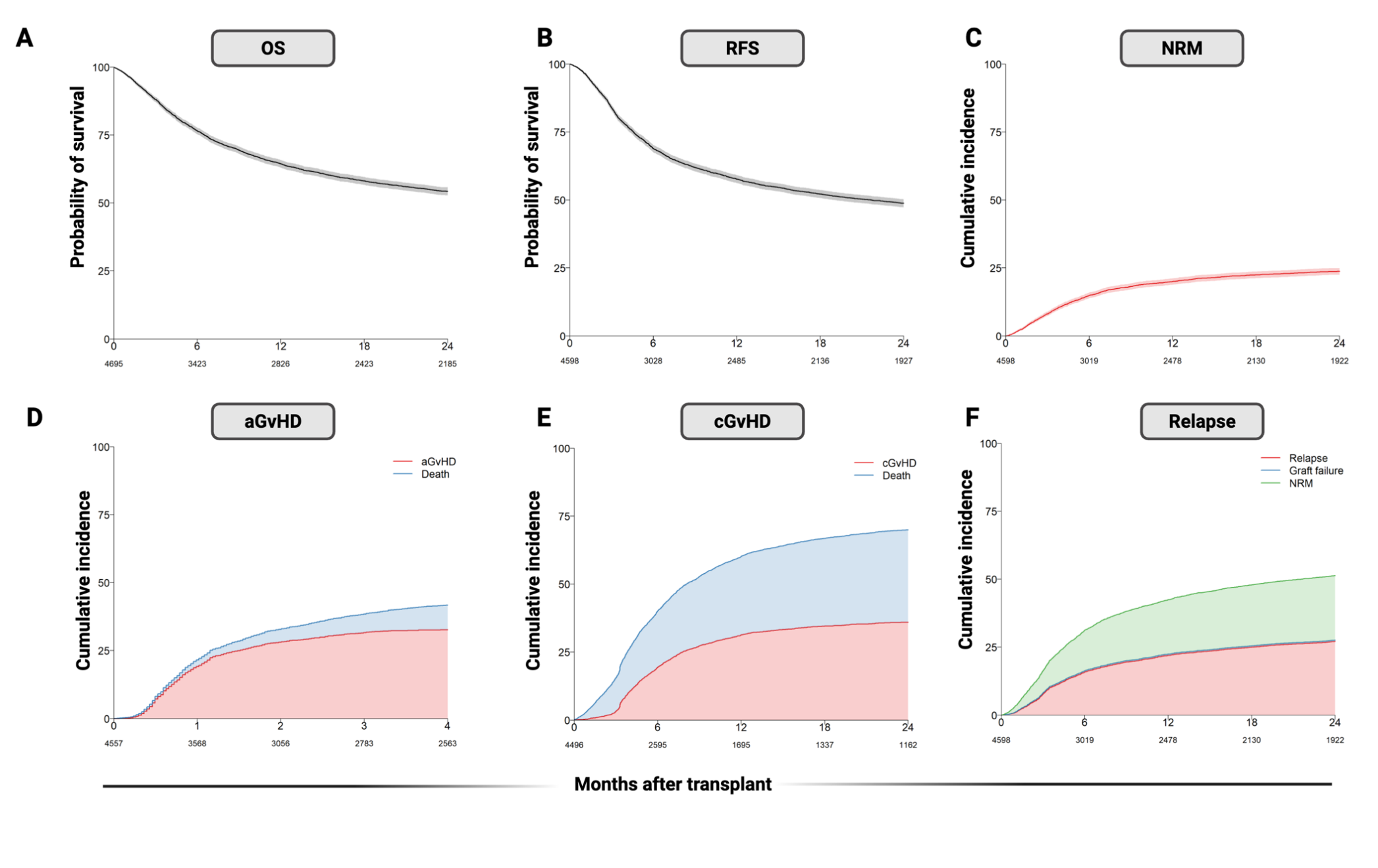

**Figure S2:** Outcomes after single HLA-mismatched unrelated donor allogeneic hematopoietic cell transplantation.

**(A–B)**, Kaplan–Meier estimates of overall survival (OS, **A**) and relapse-free survival (RFS, **B**); shaded bands indicate 95% confidence intervals.
(**C–F**), Cumulative incidence functions accounting for competing risks: (**C**) non-relapse mortality (NRM; relapse/progression treated as a competing event); (**D**) grade II–IV acute graft-versus-host disease (aGvHD) by day +100 with death before day +100 as competing risk (red area = aGvHD; blue = death without aGvHD); (**E**) chronic GvHD with death as competing risk (red = cGvHD; blue = death without cGvHD); (**F**) relapse with graft failure and NRM as competing events (red = relapse; blue = graft failure; green = NRM).
Time is shown in months after transplant; numbers below the x-axis denote patients at risk at each time point (n varies by panel due to data availability).

#### Supplementary tables

| **Table S1: univariable analysis of all the outcomes according to the mismatch type** | | | |
| --- | --- | --- | --- |
| **Outcome** | **Mismatch Type** | **2y-Survival or Cumulative Incidence (%)** | **95% CI** |
| **NRM** | A | 26.6 | (24.3 : 28.8) |
|  | B | 27.3 | (24.0 : 30.5) |
|  | C | 22.7 | (20.1 : 25.3) |
|  | DRB1 | 19.2 | (16.4 : 22.1) |
|  | DQB1 | 17.9 | (14.2 : 21.5) |
| **OS** | A | 50.7 | (48.2 : 53.3) |
|  | B | 53.0 | (49.3 : 56.6) |
|  | C | 53.7 | (50.6 : 56.7) |
|  | DRB1 | 60.5 | (57.0 : 64.0) |
|  | DQB1 | 60.2 | (55.4 : 64.9) |
| **RFS** | A | 45.0 | (42.5 : 47.5) |
|  | B | 48.4 | (44.7 : 52.1) |
|  | C | 49.1 | (46.0 : 52.2) |
|  | DRB1 | 54.1 | (50.5 : 57.7) |
|  | DQB1 | 53.0 | (48.2 : 57.9) |
| **Relapse** | A | 27.8 | (25.5 : 30.0) |
|  | B | 24.1 | (20.9 : 27.2) |
|  | C | 28.0 | (25.2 : 30.8) |
|  | DRB1 | 26.7 | (23.5 : 29.9) |
|  | DQB1 | 28.4 | (24.0 : 32.8) |
| **aGVHD*** | A | 33.9 | (31.5 : 36.2) |
|  | B | 34.8 | (31.3 : 38.2) |
|  | C | 33.7 | (30.9 : 36.6) |
|  | DRB1 | 27.1 | (23.9 : 30.2) |
|  | DQB1 | 32.3 | (27.8 : 36.8) |
| **cGVHD** | A | 35.0 | (32.5 : 37.4) |
|  | B | 35.4 | (31.9 : 38.9) |
|  | C | 39.3 | (36.3 : 42.3) |
|  | DRB1 | 33.6 | (30.2 : 37.1) |
|  | DQB1 | 36.6 | (31.9 : 41.3) |

***aGvHD incidence at 4 months**

##### **Table S2: Multivariable analysis of all the outcomes according to the mismatch type**

|  | | **OS (N=4199)** | | | **RFS (N=4119)** | | | **NRM (N=4119)** | | | **aGvHD (N=4092)** | | | **cGvHD (N=4034)** | | | **Relapse (N=4119)** | | |
| --- | --- | --- | --- | --- | --- | --- | --- | --- | --- | --- | --- | --- | --- | --- | --- | --- | --- | --- | --- |
| **Variable** | **Category** | **HR** | **95% CI** | **p-value** | **HR** | **95% CI** | **p-value** | **HR** | **95% CI** | **p-value** | **HR** | **95% CI** | **p-value** | **HR** | **95% CI** | **p-value** | **HR** | **95% CI** | **p-value** |
| **Stem-cell source** | BM (ref) | -1 |  |  | -1 |  |  | - |  |  | - |  |  | - |  |  | - |  |  |
|  | PB | 1,02 | 0.88–1.19 | 0.8 | 1,02 | 0.88–1.18 | 0.8 | 0,86 | 0.69–1.06 | 0.15 | 1,14 | 0.94–1.37 | 0.2 | 1,35 | 1.12–1.63 | 0.002 | 1,21 | 0.98–1.49 | 0.081 |
| **Patient age (per decade)** |  | 1,17 | 1.12–1.21 | <0.001 | 1,11 | 1.07–1.15 | <0.001 | 1,28 | 1.21–1.35 | <0.001 | 0,97 | 0.93–1.01 | 0.2 | 1,05 | 1.01–1.10 | 0.018 | 0,99 | 0.95–1.04 | 0.8 |
| **Donor age (per decade)** |  | 1,05 | 1.00–1.11 | 0.040 | 1,06 | 1.01–1.12 | 0.012 | 1,09 | 1.02–1.17 | 0.012 | 1,05 | 0.98–1.11 | 0.15 | 1,07 | 1.00–1.13 | 0.037 | 1,04 | 0.97–1.11 | 0.3 |
| **Disease Risk Index** | Intermediate–Low (ref) | - |  |  | - |  |  | - |  |  | - |  |  | - |  |  | - |  |  |
|  | High–Very High | 0,62 | 0.57–0.68 | <0.001 | 0,63 | 0.57–0.68 | <0.001 | 0,81 | 0.70–0.92 | 0.001 | 0,93 | 0.83–1.04 | 0.2 | 0,9 | 0.80–1.01 | 0.065 | 0,51 | 0.45–0.57 | <0.001 |
| **T-cell depletion** | No (ref) | - |  |  | - |  |  | - |  |  | - |  |  | - |  |  | - |  |  |
|  | Yes | 0,92 | 0.80–1.05 | 0.2 | 0,94 | 0.83–1.08 | 0.4 | 0,93 | 0.77–1.13 | 0.5 | 0,71 | 0.61–0.83 | <0.001 | 0,7 | 0.60–0.82 | <0.001 | 0,96 | 0.80–1.16 | 0.7 |
| **Donor sex match** | Other (ref) | - |  |  | - |  |  | - |  |  | - |  |  | - |  |  | - |  |  |
|  | Male–Female | 1,11 | 0.99–1.24 | 0.081 | 1,09 | 0.98–1.22 | 0.12 | 1,18 | 1.01–1.39 | 0.039 | 1,05 | 0.91–1.22 | 0.5 | 1,19 | 1.03–1.36 | 0.015 | 1 | 0.85–1.16 | >0.9 |
| **Karnofsky score** | ≥90 (ref) | - |  |  | - |  |  | - |  |  | - |  |  | - |  |  | - |  |  |
|  | <90 | 1,31 | 1.18–1.44 | <0.001 | 1,25 | 1.13–1.37 | <0.001 | 1,36 | 1.18–1.56 | <0.001 | 1,04 | 0.92–1.18 | 0.5 | 0,95 | 0.83–1.07 | 0.4 | 1,17 | 1.02–1.33 | 0.024 |
|  | Missing | 1,39 | 1.16–1.65 | <0.001 | 1,29 | 1.09–1.54 | 0.003 | 1,23 | 0.95–1.59 | 0.12 | 0,92 | 0.72–1.17 | 0.5 | 0,99 | 0.79–1.25 | >0.9 | 1,32 | 1.04–1.67 | 0.021 |
| **CMV serostatus** | Other (ref) | - |  |  | - |  |  | - |  |  | - |  |  | - |  |  | - |  |  |
|  | -/- | 0,81 | 0.72–0.90 | <0.001 | 0,83 | 0.75–0.92 | <0.001 | 0,78 | 0.67–0.91 | 0.001 | 1,02 | 0.90–1.16 | 0.7 | 1,01 | 0.89–1.14 | >0.9 | 0,88 | 0.77–1.01 | 0.080 |
| **Year of transplant** |  | 0,98 | 0.96–1.00 | 0.017 | 0,99 | 0.97–1.00 | 0.15 | 0,98 | 0.96–1.01 | 0.2 | 0,98 | 0.95–1.00 | 0.034 | 0,97 | 0.95–0.99 | 0.002 | 0,99 | 0.96–1.01 | 0.3 |
| **Total body irradiation** | No (ref) | - |  |  | - |  |  | - |  |  | - |  |  | - |  |  | - |  |  |
|  | Yes | 1,04 | 0.93–1.17 | 0.5 | 1 | 0.89–1.11 | >0.9 | 1,15 | 0.98–1.35 | 0.089 | 1,04 | 0.92–1.19 | 0.5 | 0,99 | 0.87–1.13 | 0.9 | 0,89 | 0.77–1.03 | 0.12 |
| **Conditioning intensity** | Standard (ref) | - |  |  | - |  |  | - |  |  | - |  |  | - |  |  | - |  |  |
|  | Reduced | 0,92 | 0.83–1.02 | 0.12 | 0,97 | 0.88–1.07 | 0.5 | 0,89 | 0.77–1.04 | 0.14 | 0,81 | 0.71–0.92 | 0.001 | 0,94 | 0.83–1.06 | 0.3 | 1,03 | 0.90–1.18 | 0.7 |
| **GVHD prophylaxis** | CSA–MTX (ref) | - |  |  | - |  |  | - |  |  | - |  |  | - |  |  | - |  |  |
|  | CSA–MMF | 1,04 | 0.93–1.16 | 0.5 | 1,05 | 0.95–1.17 | 0.3 | 1,1 | 0.94–1.29 | 0.2 | 1,22 | 1.07–1.40 | 0.003 | 1,16 | 1.02–1.32 | 0.028 |  |  |  |
|  | Tac-based | 0,94 | 0.81–1.09 | 0.4 | 0,91 | 0.79–1.04 | 0.2 | 1,06 | 0.86–1.29 | 0.6 | 1,07 | 0.90–1.27 | 0.5 | 0,96 | 0.81–1.14 | 0.6 | 0,79 | 0.65–0.96 | 0.019 |
|  | Other | 1,29 | 1.13–1.46 | <0.001 | 1,28 | 1.13–1.44 | <0.001 | 1,5 | 1.26–1.79 | <0.001 | 1,15 | 0.98–1.36 | 0.088 | 1,21 | 1.04–1.41 | 0.015 | 1,09 | 0.92–1.30 | 0.3 |
| **Mismatch** | A locus | 1,41 | 1.24–1.62 | **<0.001** | 1,32 | 1.16–1.50 | **<0.001** | 1,62 | 1.33–1.97 | **<0.001** | 1,32 | 1.11–1.56 | **0.001** | 1,23 | 1.05–1.43 | **0.010** | 1,11 | 0.94–1.31 | 0.2 |
|  | B locus | 1,38 | 1.18–1.61 | **<0.001** | 1,27 | 1.10–1.48 | **0.001** | 1,72 | 1.38–2.15 | **<0.001** | 1,36 | 1.12–1.64 | **0.002** | 1,16 | 0.97–1.39 | 0.11 | 0,98 | 0.80–1.21 | 0.9 |
|  | C locus | 1,3 | 1.13–1.50 | **<0.001** | 1,22 | 1.06–1.39 | **0.005** | 1,39 | 1.13–1.72 | **0.002** | 1,28 | 1.07–1.53 | **0.007** | 1,28 | 1.09–1.51 | **0.003** | 1,09 | 0.91–1.30 | 0.4 |
|  | DRB1 locus | 1,04 | 0.86–1.27 | 0.7 | 1,03 | 0.86–1.23 | 0.8 | 1,02 | 0.76–1.37 | 0.9 | 1,18 | 0.94–1.49 | 0.2 | 1,11 | 0.90–1.37 | 0.3 | 1 | 0.79–1.26 | >0.9 |
|  | DQB1 locus (ref) | - |  |  | - |  |  | - |  |  | - |  |  | - |  |  | - |  |  |

##### **Table S3: Multivariable Cox regression analysis of HED metrics according to mismatch group**

### Mismatch A Group (N=1382)

|  | **RFS** | | | **Relapse** | | | **NRM** | | | **OS** | | | **aGVHD** | | | **cGVHD** | | |
| --- | --- | --- | --- | --- | --- | --- | --- | --- | --- | --- | --- | --- | --- | --- | --- | --- | --- | --- |
| **Variable** | **HR** | **95% CI** | **p** | **HR** | **95% CI** | **p** | **HR** | **95% CI** | **p** | **HR** | **95% CI** | **p** | **HR** | **95% CI** | **p** | **HR** | **95% CI** | **p** |
| **Stem-cell source** |  |  |  |  |  |  |  |  |  |  |  |  |  |  |  |  |  |  |
| BM | — | — |  | — | — |  | — | — |  | — | — |  | — | — |  | — | — |  |
| PB | 0.94 | 0.73, 1.21 | 0.6 | 1.09 | 0.76, 1.55 | 0.6 | 0.81 | 0.57, 1.15 | 0.2 | 0.89 | 0.69, 1.15 | 0.4 | 0.97 | 0.71, 1.34 | 0.9 | 1.59 | 1.11, 2.29 | 0.011 |
| Patient age (decades) | 1.09 | 1.03, 1.16 | 0.004 | 0.95 | 0.87, 1.03 | 0.2 | 1.29 | 1.18, 1.42 | <0.001 | 1.14 | 1.07, 1.22 | <0.001 | 0.94 | 0.87, 1.01 | 0.078 | 1.02 | 0.94, 1.10 | 0.6 |
| Donor age (decades) | 1.05 | 0.96, 1.14 | 0.3 | 1.04 | 0.92, 1.17 | 0.5 | 1.07 | 0.95, 1.20 | 0.3 | 1.02 | 0.94, 1.12 | 0.6 | 0.99 | 0.88, 1.10 | 0.8 | 1.09 | 0.98, 1.21 | 0.12 |
| **DRI** |  |  |  |  |  |  |  |  |  |  |  |  |  |  |  |  |  |  |
| Intermediate-Low | — | — |  | — | — |  | — | — |  | — | — |  | — | — |  | — | — |  |
| High-Very High | 0.72 | 0.62, 0.83 | <0.001 | 0.54 | 0.44, 0.66 | <0.001 | 1.00 | 0.79, 1.25 | >0.9 | 0.71 | 0.61, 0.83 | <0.001 | 0.84 | 0.69, 1.02 | 0.086 | 0.97 | 0.79, 1.19 | 0.8 |
| **T-cell depletion** |  |  |  |  |  |  |  |  |  |  |  |  |  |  |  |  |  |  |
| No | — | — |  | — | — |  | — | — |  | — | — |  | — | — |  | — | — |  |
| Yes | 0.97 | 0.75, 1.26 | 0.8 | 0.76 | 0.54, 1.07 | 0.12 | 1.31 | 0.87, 1.96 | 0.2 | 0.98 | 0.75, 1.27 | 0.9 | 0.90 | 0.66, 1.23 | 0.5 | 0.67 | 0.50, 0.90 | 0.007 |
| **Donor sex match** |  |  |  |  |  |  |  |  |  |  |  |  |  |  |  |  |  |  |
| Other | — | — |  | — | — |  | — | — |  | — | — |  | — | — |  | — | — |  |
| Male-female | 1.29 | 1.07, 1.54 | 0.006 | 1.27 | 0.99, 1.64 | 0.063 | 1.27 | 0.98, 1.64 | 0.066 | 1.34 | 1.11, 1.60 | 0.002 | 1.06 | 0.83, 1.35 | 0.6 | 1.09 | 0.85, 1.39 | 0.5 |
| **Karnofsky score** |  |  |  |  |  |  |  |  |  |  |  |  |  |  |  |  |  |  |
| ≥90 | — | — |  | — | — |  | — | — |  | — | — |  | — | — |  | — | — |  |
| <90 | 1.28 | 1.09, 1.50 | 0.003 | 1.20 | 0.95, 1.51 | 0.12 | 1.39 | 1.11, 1.74 | 0.004 | 1.27 | 1.08, 1.49 | 0.005 | 0.88 | 0.71, 1.10 | 0.3 | 1.05 | 0.85, 1.29 | 0.7 |
| **CMV -/- vs. other** |  |  |  |  |  |  |  |  |  |  |  |  |  |  |  |  |  |  |
| Other combination | — | — |  | — | — |  | — | — |  | — | — |  | — | — |  | — | — |  |
| -/- | 0.81 | 0.68, 0.97 | 0.022 | 0.78 | 0.60, 1.00 | 0.046 | 0.86 | 0.67, 1.11 | 0.2 | 0.80 | 0.67, 0.97 | 0.020 | 0.95 | 0.76, 1.18 | 0.6 | 0.99 | 0.80, 1.22 | >0.9 |
| Year transplant | 0.98 | 0.95, 1.02 | 0.3 | 0.97 | 0.93, 1.02 | 0.2 | 1.00 | 0.95, 1.04 | 0.9 | 0.98 | 0.94, 1.01 | 0.13 | 0.96 | 0.93, 1.00 | 0.062 | 0.95 | 0.91, 0.99 | 0.010 |
| **Total body irradiation** |  |  |  |  |  |  |  |  |  |  |  |  |  |  |  |  |  |  |
| No | — | — |  | — | — |  | — | — |  | — | — |  | — | — |  | — | — |  |
| Yes | 0.95 | 0.79, 1.14 | 0.6 | 0.79 | 0.61, 1.02 | 0.075 | 1.15 | 0.88, 1.51 | 0.3 | 1.01 | 0.83, 1.22 | >0.9 | 1.03 | 0.83, 1.29 | 0.8 | 0.96 | 0.76, 1.21 | 0.7 |
| **Conditioning intensity** |  |  |  |  |  |  |  |  |  |  |  |  |  |  |  |  |  |  |
| Standard | — | — |  | — | — |  | — | — |  | — | — |  | — | — |  | — | — |  |
| Reduced | 0.98 | 0.82, 1.16 | 0.8 | 1.05 | 0.82, 1.34 | 0.7 | 0.88 | 0.69, 1.12 | 0.3 | 0.98 | 0.82, 1.17 | 0.8 | 0.87 | 0.70, 1.08 | 0.2 | 0.90 | 0.72, 1.12 | 0.3 |
| **GVHD prophylaxis** |  |  |  |  |  |  |  |  |  |  |  |  |  |  |  |  |  |  |
| CSA-MTX | — | — |  | — | — |  | — | — |  | — | — |  | — | — |  | — | — |  |
| CSA-MMF | 1.01 | 0.84, 1.22 | 0.9 | 0.97 | 0.75, 1.25 | 0.8 | 1.10 | 0.84, 1.45 | 0.5 | 1.11 | 0.92, 1.34 | 0.3 | 1.47 | 1.16, 1.86 | 0.001 | 1.33 | 1.05, 1.68 | 0.017 |
| Tac-based | 0.93 | 0.74, 1.18 | 0.6 | 0.72 | 0.51, 1.01 | 0.059 | 1.22 | 0.89, 1.68 | 0.2 | 1.01 | 0.80, 1.29 | >0.9 | 1.43 | 1.08, 1.89 | 0.013 | 1.04 | 0.77, 1.40 | 0.8 |
| Other | 1.16 | 0.94, 1.42 | 0.2 | 0.92 | 0.69, 1.25 | 0.6 | 1.43 | 1.07, 1.90 | 0.014 | 1.22 | 0.99, 1.50 | 0.068 | 1.25 | 0.95, 1.64 | 0.11 | 1.36 | 1.05, 1.76 | 0.021 |
| A_HED-R | 1.00 | 0.97, 1.02 | 0.7 | 0.99 | 0.95, 1.02 | 0.4 | 1.01 | 0.98, 1.04 | 0.6 | 1.00 | 0.97, 1.02 | 0.8 | 1.02 | 0.99, 1.05 | 0.13 | 1.03 | 1.00, 1.06 | **0.044** |
| B_HED-R | 1.02 | 0.99, 1.04 | 0.13 | 1.01 | 0.98, 1.05 | 0.4 | 1.02 | 0.99, 1.06 | 0.2 | 1.01 | 0.98, 1.03 | 0.5 | 1.00 | 0.97, 1.02 | 0.8 | 1.00 | 0.97, 1.03 | 0.8 |
| C_HED-R | 1.00 | 0.96, 1.03 | 0.8 | 1.00 | 0.95, 1.04 | 0.8 | 1.00 | 0.95, 1.05 | >0.9 | 1.00 | 0.97, 1.04 | 0.9 | 0.99 | 0.94, 1.03 | 0.5 | 1.03 | 0.99, 1.08 | 0.2 |
| DRB1_HED-R | 1.00 | 0.98, 1.01 | 0.9 | 1.00 | 0.98, 1.02 | >0.9 | 1.00 | 0.97, 1.02 | 0.8 | 1.00 | 0.98, 1.02 | >0.9 | 1.01 | 0.99, 1.03 | 0.4 | 0.99 | 0.97, 1.01 | 0.2 |
| DQB1_HED-R | 0.99 | 0.98, 1.01 | 0.3 | 0.99 | 0.97, 1.01 | 0.2 | 1.00 | 0.98, 1.01 | 0.7 | 1.00 | 0.98, 1.01 | 0.4 | 0.99 | 0.98, 1.01 | 0.4 | 1.00 | 0.98, 1.01 | 0.9 |
| HED-MM A | 1.00 | 0.97, 1.02 | 0.9 | 1.00 | 0.96, 1.03 | 0.8 | 1.00 | 0.97, 1.04 | 0.9 | 1.00 | 0.97, 1.02 | 0.8 | 0.99 | 0.96, 1.02 | 0.5 | 1.02 | 0.98, 1.05 | 0.4 |

Mismatch B Group (N=639)

|  | **RFS** | | | **Relapse** | | | **NRM** | | | **OS** | | | **aGVHD** | | | **cGVHD** | | |
| --- | --- | --- | --- | --- | --- | --- | --- | --- | --- | --- | --- | --- | --- | --- | --- | --- | --- | --- |
| **Variable** | **HR** | **95% CI** | **p** | **HR** | **95% CI** | **p** | **HR** | **95% CI** | **p** | **HR** | **95% CI** | **p** | **HR** | **95% CI** | **p** | **HR** | **95% CI** | **p** |
| **Stem-cell source** |  |  |  |  |  |  |  |  |  |  |  |  |  |  |  |  |  |  |
| BM | — | — |  | — | — |  | — | — |  | — | — |  | — | — |  | — | — |  |
| PB | 1.10 | 0.75, 1.60 | 0.6 | 1.43 | 0.79, 2.58 | 0.2 | 0.89 | 0.54, 1.46 | 0.6 | 1.22 | 0.82, 1.81 | 0.3 | 1.20 | 0.75, 1.93 | 0.5 | 1.33 | 0.82, 2.17 | 0.2 |
| Patient age (decades) | 1.05 | 0.96, 1.14 | 0.3 | 0.86 | 0.77, 0.97 | 0.014 | 1.29 | 1.13, 1.47 | <0.001 | 1.15 | 1.05, 1.26 | 0.002 | 1.01 | 0.91, 1.12 | 0.9 | 1.12 | 1.00, 1.25 | 0.045 |
| Donor age (decades) | 1.02 | 0.90, 1.15 | 0.8 | 1.09 | 0.91, 1.31 | 0.3 | 0.98 | 0.82, 1.16 | 0.8 | 1.04 | 0.91, 1.18 | 0.6 | 0.94 | 0.79, 1.10 | 0.4 | 1.14 | 0.97, 1.34 | 0.12 |
| **DRI** |  |  |  |  |  |  |  |  |  |  |  |  |  |  |  |  |  |  |
| Intermediate-Low | — | — |  | — | — |  | — | — |  | — | — |  | — | — |  | — | — |  |
| High-Very High | 0.61 | 0.49, 0.76 | <0.001 | 0.55 | 0.40, 0.75 | <0.001 | 0.70 | 0.51, 0.95 | 0.024 | 0.61 | 0.49, 0.77 | <0.001 | 0.95 | 0.72, 1.26 | 0.7 | 0.61 | 0.46, 0.81 | <0.001 |
| **T-cell depletion** |  |  |  |  |  |  |  |  |  |  |  |  |  |  |  |  |  |  |
| No | — | — |  | — | — |  | — | — |  | — | — |  | — | — |  | — | — |  |
| Yes | 0.91 | 0.66, 1.25 | 0.6 | 1.03 | 0.64, 1.66 | >0.9 | 0.80 | 0.52, 1.25 | 0.3 | 0.88 | 0.63, 1.22 | 0.4 | 0.85 | 0.57, 1.26 | 0.4 | 1.07 | 0.71, 1.63 | 0.7 |
| **Donor sex match** |  |  |  |  |  |  |  |  |  |  |  |  |  |  |  |  |  |  |
| Other | — | — |  | — | — |  | — | — |  | — | — |  | — | — |  | — | — |  |
| Male-female | 1.43 | 1.09, 1.87 | 0.009 | 1.08 | 0.71, 1.66 | 0.7 | 1.72 | 1.21, 2.47 | 0.003 | 1.48 | 1.12, 1.94 | 0.006 | 0.85 | 0.57, 1.25 | 0.4 | 0.93 | 0.63, 1.38 | 0.7 |
| **Karnofsky score** |  |  |  |  |  |  |  |  |  |  |  |  |  |  |  |  |  |  |
| ≥90 | — | — |  | — | — |  | — | — |  | — | — |  | — | — |  | — | — |  |
| <90 | 1.21 | 0.95, 1.56 | 0.13 | 1.19 | 0.83, 1.71 | 0.3 | 1.23 | 0.87, 1.75 | 0.2 | 1.30 | 1.01, 1.68 | 0.043 | 0.99 | 0.71, 1.38 | >0.9 | 0.74 | 0.52, 1.05 | 0.094 |
| **CMV -/- vs. other** |  |  |  |  |  |  |  |  |  |  |  |  |  |  |  |  |  |  |
| Other combination | — | — |  | — | — |  | — | — |  | — | — |  | — | — |  | — | — |  |
| -/- | 0.74 | 0.57, 0.96 | 0.023 | 0.80 | 0.55, 1.16 | 0.2 | 0.68 | 0.47, 0.99 | 0.043 | 0.77 | 0.59, 1.01 | 0.057 | 0.82 | 0.59, 1.13 | 0.2 | 1.01 | 0.74, 1.37 | >0.9 |
| Year transplant | 1.01 | 0.96, 1.06 | 0.6 | 1.01 | 0.94, 1.08 | 0.9 | 1.01 | 0.94, 1.07 | 0.9 | 1.00 | 0.95, 1.05 | >0.9 | 1.01 | 0.95, 1.08 | 0.7 | 0.94 | 0.89, 1.00 | 0.045 |
| **Total body irradiation** |  |  |  |  |  |  |  |  |  |  |  |  |  |  |  |  |  |  |
| No | — | — |  | — | — |  | — | — |  | — | — |  | — | — |  | — | — |  |
| Yes | 1.19 | 0.91, 1.56 | 0.2 | 0.97 | 0.66, 1.43 | 0.9 | 1.45 | 0.99, 2.11 | 0.054 | 1.23 | 0.94, 1.62 | 0.14 | 1.01 | 0.73, 1.41 | >0.9 | 1.27 | 0.91, 1.77 | 0.2 |
| **Conditioning intensity** |  |  |  |  |  |  |  |  |  |  |  |  |  |  |  |  |  |  |
| Standard | — | — |  | — | — |  | — | — |  | — | — |  | — | — |  | — | — |  |
| Reduced | 1.16 | 0.89, 1.50 | 0.3 | 1.19 | 0.82, 1.74 | 0.4 | 1.13 | 0.79, 1.62 | 0.5 | 0.97 | 0.74, 1.27 | 0.8 | 0.89 | 0.65, 1.22 | 0.5 | 1.00 | 0.72, 1.40 | >0.9 |
| **GVHD prophylaxis** |  |  |  |  |  |  |  |  |  |  |  |  |  |  |  |  |  |  |
| CSA-MTX | — | — |  | — | — |  | — | — |  | — | — |  | — | — |  | — | — |  |
| CSA-MMF | 1.12 | 0.86, 1.46 | 0.4 | 1.21 | 0.82, 1.78 | 0.3 | 1.05 | 0.73, 1.52 | 0.8 | 0.97 | 0.74, 1.29 | 0.8 | 0.98 | 0.70, 1.36 | >0.9 | 1.10 | 0.79, 1.54 | 0.6 |
| Tac-based | 0.92 | 0.64, 1.31 | 0.6 | 1.02 | 0.62, 1.67 | >0.9 | 0.78 | 0.46, 1.34 | 0.4 | 0.88 | 0.60, 1.28 | 0.5 | 1.03 | 0.68, 1.56 | 0.9 | 0.95 | 0.61, 1.47 | 0.8 |
| Other | 1.31 | 0.96, 1.79 | 0.090 | 1.31 | 0.82, 2.09 | 0.3 | 1.30 | 0.85, 2.00 | 0.2 | 1.22 | 0.88, 1.69 | 0.2 | 0.79 | 0.51, 1.23 | 0.3 | 0.71 | 0.46, 1.10 | 0.13 |
| A_HED-R | 1.02 | 0.99, 1.05 | 0.13 | 1.01 | 0.97, 1.05 | 0.5 | 1.03 | 0.99, 1.07 | 0.14 | 1.02 | 0.99, 1.05 | 0.2 | 1.01 | 0.98, 1.05 | 0.6 | 0.97 | 0.94, 1.01 | 0.13 |
| B_HED-R | 1.01 | 0.98, 1.05 | 0.6 | 0.99 | 0.95, 1.04 | 0.8 | 1.02 | 0.98, 1.07 | 0.3 | 1.01 | 0.98, 1.05 | 0.5 | 0.99 | 0.95, 1.04 | 0.8 | 0.99 | 0.95, 1.04 | 0.8 |
| C_HED-R | 1.02 | 0.97, 1.07 | 0.5 | 1.00 | 0.93, 1.07 | >0.9 | 1.03 | 0.96, 1.10 | 0.5 | 1.04 | 0.98, 1.09 | 0.2 | 1.02 | 0.95, 1.08 | 0.6 | 1.00 | 0.94, 1.06 | >0.9 |
| DRB1_HED-R | 1.03 | 1.01, 1.05 | **0.016** | 1.03 | 1.00, 1.07 | **0.046** | 1.02 | 0.99, 1.06 | 0.14 | 1.03 | 1.01, 1.06 | **0.007** | 1.00 | 0.97, 1.03 | 0.8 | 1.01 | 0.98, 1.03 | 0.7 |
| DQB1_HED-R | 0.98 | 0.96, 1.00 | **0.046** | 0.98 | 0.96, 1.01 | 0.2 | 0.98 | 0.96, 1.00 | 0.10 | 0.98 | 0.96, 1.00 | **0.027** | 1.02 | 0.99, 1.04 | 0.2 | 0.99 | 0.97, 1.01 | 0.4 |
| HED-MM B | 1.01 | 0.98, 1.04 | 0.6 | 1.01 | 0.97, 1.07 | 0.6 | 1.01 | 0.96, 1.06 | 0.7 | 1.01 | 0.98, 1.05 | 0.5 | 0.99 | 0.95, 1.03 | 0.7 | 0.96 | 0.92, 1.01 | 0.11 |

Mismatch C Group (N=937)

|  | **RFS** | | | **Relapse** | | | **NRM** | | | **OS** | | | **aGVHD** | | | **cGVHD** | | |
| --- | --- | --- | --- | --- | --- | --- | --- | --- | --- | --- | --- | --- | --- | --- | --- | --- | --- | --- |
| **Variable** | **HR** | **95% CI** | **p** | **HR** | **95% CI** | **p** | **HR** | **95% CI** | **p** | **HR** | **95% CI** | **p** | **HR** | **95% CI** | **p** | **HR** | **95% CI** | **p** |
| **Stem-cell source** |  |  |  |  |  |  |  |  |  |  |  |  |  |  |  |  |  |  |
| BM | — | — |  | — | — |  | — | — |  | — | — |  | — | — |  | — | — |  |
| PB | 0.93 | 0.69, 1.24 | 0.6 | 1.06 | 0.70, 1.61 | 0.8 | 0.78 | 0.51, 1.20 | 0.3 | 0.99 | 0.73, 1.36 | >0.9 | 1.31 | 0.88, 1.94 | 0.2 | 1.38 | 0.95, 2.00 | 0.086 |
| Patient age (decades) | 1.20 | 1.11, 1.30 | <0.001 | 1.16 | 1.04, 1.29 | 0.005 | 1.27 | 1.13, 1.43 | <0.001 | 1.22 | 1.13, 1.33 | <0.001 | 0.96 | 0.88, 1.05 | 0.3 | 1.05 | 0.96, 1.15 | 0.3 |
| Donor age (decades) | 1.10 | 1.00, 1.22 | 0.048 | 1.01 | 0.88, 1.15 | >0.9 | 1.24 | 1.07, 1.43 | 0.005 | 1.13 | 1.02, 1.25 | 0.021 | 1.11 | 0.98, 1.25 | 0.10 | 1.10 | 0.98, 1.23 | 0.10 |
| **DRI** |  |  |  |  |  |  |  |  |  |  |  |  |  |  |  |  |  |  |
| Intermediate-Low | — | — |  | — | — |  | — | — |  | — | — |  | — | — |  | — | — |  |
| High-Very High | 0.57 | 0.47, 0.69 | <0.001 | 0.46 | 0.36, 0.59 | <0.001 | 0.75 | 0.56, 1.00 | 0.049 | 0.57 | 0.47, 0.69 | <0.001 | 0.93 | 0.73, 1.18 | 0.5 | 0.75 | 0.60, 0.95 | 0.017 |
| **T-cell depletion** |  |  |  |  |  |  |  |  |  |  |  |  |  |  |  |  |  |  |
| No | — | — |  | — | — |  | — | — |  | — | — |  | — | — |  | — | — |  |
| Yes | 1.00 | 0.74, 1.34 | >0.9 | 1.05 | 0.69, 1.60 | 0.8 | 0.96 | 0.63, 1.47 | 0.9 | 1.02 | 0.75, 1.39 | >0.9 | 0.67 | 0.48, 0.93 | 0.017 | 0.62 | 0.45, 0.86 | 0.004 |
| **Donor sex match** |  |  |  |  |  |  |  |  |  |  |  |  |  |  |  |  |  |  |
| Other | — | — |  | — | — |  | — | — |  | — | — |  | — | — |  | — | — |  |
| Male-female | 0.80 | 0.63, 1.02 | 0.068 | 0.82 | 0.59, 1.14 | 0.2 | 0.76 | 0.53, 1.10 | 0.15 | 0.74 | 0.57, 0.96 | 0.021 | 1.19 | 0.89, 1.58 | 0.2 | 1.07 | 0.82, 1.41 | 0.6 |
| **Karnofsky score** |  |  |  |  |  |  |  |  |  |  |  |  |  |  |  |  |  |  |
| ≥90 | — | — |  | — | — |  | — | — |  | — | — |  | — | — |  | — | — |  |
| <90 | 1.27 | 1.03, 1.57 | 0.027 | 1.06 | 0.79, 1.43 | 0.7 | 1.57 | 1.16, 2.13 | 0.004 | 1.35 | 1.09, 1.67 | 0.007 | 1.35 | 1.04, 1.74 | 0.022 | 0.94 | 0.72, 1.23 | 0.6 |
| **CMV -/- vs. other** |  |  |  |  |  |  |  |  |  |  |  |  |  |  |  |  |  |  |
| Other combination | — | — |  | — | — |  | — | — |  | — | — |  | — | — |  | — | — |  |
| -/- | 0.85 | 0.69, 1.05 | 0.13 | 0.92 | 0.70, 1.21 | 0.5 | 0.75 | 0.54, 1.03 | 0.079 | 0.77 | 0.62, 0.96 | 0.020 | 1.25 | 0.98, 1.60 | 0.077 | 0.91 | 0.71, 1.16 | 0.5 |
| Year transplant | 0.99 | 0.96, 1.03 | 0.7 | 1.01 | 0.96, 1.06 | 0.7 | 0.97 | 0.92, 1.03 | 0.3 | 0.99 | 0.95, 1.03 | 0.6 | 0.99 | 0.94, 1.03 | 0.6 | 0.98 | 0.94, 1.03 | 0.5 |
| **Total body irradiation** |  |  |  |  |  |  |  |  |  |  |  |  |  |  |  |  |  |  |
| No | — | — |  | — | — |  | — | — |  | — | — |  | — | — |  | — | — |  |
| Yes | 0.96 | 0.76, 1.20 | 0.7 | 0.81 | 0.59, 1.11 | 0.2 | 1.16 | 0.84, 1.61 | 0.4 | 1.07 | 0.85, 1.34 | 0.6 | 1.06 | 0.82, 1.38 | 0.7 | 0.95 | 0.73, 1.24 | 0.7 |
| **Conditioning intensity** |  |  |  |  |  |  |  |  |  |  |  |  |  |  |  |  |  |  |
| Standard | — | — |  | — | — |  | — | — |  | — | — |  | — | — |  | — | — |  |
| Reduced | 0.91 | 0.73, 1.12 | 0.4 | 0.94 | 0.71, 1.24 | 0.6 | 0.86 | 0.62, 1.18 | 0.3 | 0.92 | 0.74, 1.14 | 0.4 | 0.75 | 0.58, 0.98 | 0.038 | 0.96 | 0.74, 1.24 | 0.7 |
| **GVHD prophylaxis** |  |  |  |  |  |  |  |  |  |  |  |  |  |  |  |  |  |  |
| CSA-MTX | — | — |  | — | — |  | — | — |  | — | — |  | — | — |  | — | — |  |
| CSA-MMF | 1.05 | 0.85, 1.29 | 0.7 | 0.99 | 0.75, 1.31 | >0.9 | 1.12 | 0.81, 1.55 | 0.5 | 1.04 | 0.84, 1.29 | 0.7 | 1.08 | 0.83, 1.41 | 0.6 | 0.91 | 0.71, 1.17 | 0.5 |
| Tac-based | 0.71 | 0.51, 1.00 | 0.049 | 0.41 | 0.24, 0.71 | 0.001 | 1.14 | 0.72, 1.78 | 0.6 | 0.80 | 0.57, 1.12 | 0.2 | 0.90 | 0.61, 1.32 | 0.6 | 0.75 | 0.51, 1.08 | 0.12 |
| Other | 1.44 | 1.11, 1.88 | 0.006 | 1.19 | 0.83, 1.71 | 0.3 | 1.85 | 1.26, 2.70 | 0.002 | 1.47 | 1.12, 1.92 | 0.005 | 1.07 | 0.75, 1.53 | 0.7 | 0.90 | 0.64, 1.27 | 0.6 |
| A_HED-R | 1.00 | 0.98, 1.02 | >0.9 | 1.01 | 0.98, 1.04 | 0.5 | 0.99 | 0.96, 1.02 | 0.6 | 1.00 | 0.98, 1.03 | 0.8 | 1.01 | 0.98, 1.03 | 0.7 | 0.98 | 0.96, 1.01 | 0.2 |
| B_HED-R | 0.97 | 0.95, 1.00 | 0.051 | 0.97 | 0.94, 1.01 | 0.12 | 0.98 | 0.94, 1.02 | 0.2 | 0.99 | 0.96, 1.02 | 0.4 | 0.99 | 0.96, 1.03 | 0.7 | 1.01 | 0.97, 1.04 | 0.7 |
| C_HED-R | 1.01 | 0.96, 1.06 | 0.7 | 0.98 | 0.92, 1.05 | 0.6 | 1.04 | 0.97, 1.12 | 0.3 | 1.01 | 0.97, 1.06 | 0.5 | 1.04 | 0.98, 1.10 | 0.2 | 1.03 | 0.98, 1.09 | 0.3 |
| DRB1_HED-R | 1.00 | 0.98, 1.02 | >0.9 | 1.00 | 0.97, 1.02 | 0.8 | 1.00 | 0.98, 1.03 | 0.8 | 1.00 | 0.99, 1.02 | 0.6 | 1.00 | 0.97, 1.02 | 0.9 | 1.00 | 0.98, 1.02 | >0.9 |
| DQB1_HED-R | 1.00 | 0.98, 1.01 | 0.7 | 1.00 | 0.98, 1.02 | >0.9 | 1.00 | 0.97, 1.02 | 0.8 | 1.00 | 0.98, 1.01 | 0.7 | 1.01 | 0.99, 1.03 | 0.5 | 1.01 | 1.00, 1.03 | 0.2 |
| HED-MM C | 1.01 | 0.97, 1.04 | 0.7 | 1.00 | 0.96, 1.05 | 0.8 | 1.01 | 0.96, 1.06 | 0.6 | 1.00 | 0.97, 1.04 | 0.8 | 1.03 | 0.99, 1.08 | 0.13 | 1.01 | 0.97, 1.05 | 0.6 |

Mismatch DRB1 Group (N=331)

|  | **RFS** | | | **Relapse** | | | **NRM** | | | **OS** | | | **aGVHD** | | | **cGVHD** | | |
| --- | --- | --- | --- | --- | --- | --- | --- | --- | --- | --- | --- | --- | --- | --- | --- | --- | --- | --- |
| **Variable** | **HR** | **95% CI** | **p** | **HR** | **95% CI** | **p** | **HR** | **95% CI** | **p** | **HR** | **95% CI** | **p** | **HR** | **95% CI** | **p** | **HR** | **95% CI** | **p** |
| **Stem-cell source** |  |  |  |  |  |  |  |  |  |  |  |  |  |  |  |  |  |  |
| BM | — | — |  | — | — |  | — | — |  | — | — |  | — | — |  | — | — |  |
| PB | 1.10 | 0.62–1.94 | 0.751 | 2.01 | 0.83, 4.90 | 0.12 | 0.62 | 0.27–1.39 | 0.245 | 0.95 | 0.53–1.73 | 0.878 | 1.24 | 0.61, 2.52 | 0.5 | 0.79 | 0.41, 1.50 | 0.5 |
| Patient age (decades) | 1.23 | 1.07–1.42 | 0.004 | 1.21 | 1.01, 1.45 | 0.039 | 1.31 | 1.03–1.67 | 0.029 | 1.32 | 1.13–1.53 | <0.001 | 1.08 | 0.92, 1.27 | 0.4 | 1.06 | 0.90, 1.24 | 0.5 |
| Donor age (decades) | 1.13 | 0.93–1.37 | 0.236 | 1.04 | 0.80, 1.35 | 0.8 | 1.15 | 0.84–1.57 | 0.392 | 1.06 | 0.87–1.31 | 0.554 | 1.21 | 0.96, 1.54 | 0.11 | 0.85 | 0.67, 1.08 | 0.2 |
| **DRI** |  |  |  |  |  |  |  |  |  |  |  |  |  |  |  |  |  |  |
| Intermediate-Low | — | — |  | — | — |  | — | — |  | — | — |  | — | — |  | — | — |  |
| High-Very High | 0.48 | 0.34–0.68 | <0.001 | 0.39 | 0.25, 0.60 | <0.001 | 0.61 | 0.35–1.09 | 0.095 | 0.45 | 0.32–0.65 | <0.001 | 0.96 | 0.63, 1.47 | 0.9 | 1.10 | 0.73, 1.68 | 0.6 |
| **T-cell depletion** |  |  |  |  |  |  |  |  |  |  |  |  |  |  |  |  |  |  |
| No | — | — |  | — | — |  | — | — |  | — | — |  | — | — |  | — | — |  |
| Yes | 1.06 | 0.65–1.73 | 0.824 | 1.37 | 0.69, 2.71 | 0.4 | 0.64 | 0.31–1.35 | 0.242 | 1.07 | 0.65–1.77 | 0.791 | 0.67 | 0.39, 1.13 | 0.13 | 0.66 | 0.39, 1.10 | 0.11 |
| **Donor sex match** |  |  |  |  |  |  |  |  |  |  |  |  |  |  |  |  |  |  |
| Other | — | — |  | — | — |  | — | — |  | — | — |  | — | — |  | — | — |  |
| Male-female | 0.76 | 0.43–1.33 | 0.332 | 0.42 | 0.18, 1.01 | 0.053 | 1.41 | 0.63–3.15 | 0.401 | 1.00 | 0.58–1.75 | 0.993 | 2.01 | 1.11, 3.66 | 0.022 | 2.35 | 1.40, 3.94 | 0.001 |
| **Karnofsky score** |  |  |  |  |  |  |  |  |  |  |  |  |  |  |  |  |  |  |
| ≥90 | — | — |  | — | — |  | — | — |  | — | — |  | — | — |  | — | — |  |
| <90 | 1.04 | 0.70–1.55 | 0.840 | 1.04 | 0.61, 1.78 | 0.9 | 1.19 | 0.65–2.19 | 0.575 | 1.09 | 0.73–1.65 | 0.665 | 1.02 | 0.65, 1.61 | >0.9 | 1.34 | 0.87, 2.09 | 0.2 |
| **CMV -/- vs. other** |  |  |  |  |  |  |  |  |  |  |  |  |  |  |  |  |  |  |
| Other combination | — | — |  | — | — |  | — | — |  | — | — |  | — | — |  | — | — |  |
| -/- | 0.82 | 0.54–1.23 | 0.332 | 0.73 | 0.43, 1.26 | 0.3 | 1.03 | 0.54–1.97 | 0.929 | 0.84 | 0.55–1.29 | 0.429 | 1.30 | 0.82, 2.06 | 0.3 | 1.50 | 0.97, 2.32 | 0.067 |
| Year transplant | 0.91 | 0.85–0.97 | 0.004 | 0.91 | 0.84, 0.99 | 0.030 | 0.90 | 0.81–1.01 | 0.068 | 0.91 | 0.85–0.97 | 0.007 | 0.95 | 0.87, 1.03 | 0.2 | 0.99 | 0.92, 1.07 | 0.9 |
| **Total body irradiation** |  |  |  |  |  |  |  |  |  |  |  |  |  |  |  |  |  |  |
| No | — | — |  | — | — |  | — | — |  | — | — |  | — | — |  | — | — |  |
| Yes | 1.21 | 0.79–1.86 | 0.379 | 1.92 | 1.13, 3.28 | 0.017 | 0.60 | 0.27–1.32 | 0.202 | 1.15 | 0.74–1.80 | 0.537 | 0.99 | 0.61, 1.61 | >0.9 | 1.55 | 0.99, 2.43 | 0.054 |
| **Conditioning intensity** |  |  |  |  |  |  |  |  |  |  |  |  |  |  |  |  |  |  |
| Standard | — | — |  | — | — |  | — | — |  | — | — |  | — | — |  | — | — |  |
| Reduced | 0.75 | 0.52–1.08 | 0.125 | 0.70 | 0.43, 1.14 | 0.2 | 0.89 | 0.49–1.62 | 0.696 | 0.63 | 0.42–0.93 | 0.020 | 0.64 | 0.40, 1.01 | 0.055 | 0.97 | 0.62, 1.53 | >0.9 |
| **GVHD prophylaxis** |  |  |  |  |  |  |  |  |  |  |  |  |  |  |  |  |  |  |
| CSA-MTX | — | — |  | — | — |  | — | — |  | — | — |  | — | — |  | — | — |  |
| CSA-MMF | 1.00 | 0.67–1.50 | 0.983 | 0.93 | 0.56, 1.55 | 0.8 | 1.22 | 0.61–2.43 | 0.568 | 1.01 | 0.65–1.55 | 0.975 | 1.67 | 1.00, 2.80 | 0.051 | 1.36 | 0.83, 2.22 | 0.2 |
| Tac-based | 0.77 | 0.45–1.32 | 0.338 | 0.68 | 0.34, 1.38 | 0.3 | 0.99 | 0.41–2.36 | 0.980 | 0.80 | 0.45–1.40 | 0.431 | 0.89 | 0.46, 1.70 | 0.7 | 1.27 | 0.72, 2.24 | 0.4 |
| Other | 1.19 | 0.73–1.95 | 0.489 | 0.70 | 0.33, 1.49 | 0.4 | 2.00 | 0.95–4.21 | 0.066 | 1.34 | 0.80–2.23 | 0.264 | 1.62 | 0.92, 2.83 | 0.092 | 1.87 | 1.07, 3.28 | 0.028 |
| A_HED-R | 0.99 | 0.95–1.04 | 0.666 | 0.95 | 0.90, 1.01 | 0.10 | 1.05 | 0.97–1.13 | 0.203 | 0.99 | 0.95–1.04 | 0.834 | 1.07 | 1.01, 1.13 | **0.015** | 0.99 | 0.94, 1.04 | 0.7 |
| B_HED-R | 0.98 | 0.93–1.04 | 0.519 | 0.95 | 0.88, 1.03 | 0.2 | 1.02 | 0.93–1.12 | 0.648 | 0.98 | 0.92–1.04 | 0.471 | 0.99 | 0.93, 1.06 | 0.8 | 0.94 | 0.88, 1.00 | 0.061 |
| C_HED-R | 0.98 | 0.90–1.06 | 0.560 | 0.98 | 0.88, 1.09 | 0.7 | 1.32 | 1.00–1.73 | **0.047** | 0.97 | 0.89–1.06 | 0.494 | 1.06 | 0.96, 1.17 | 0.2 | 0.97 | 0.89, 1.07 | 0.6 |
| DRB1_HED-R | 1.04 | 1.00–1.08 | **0.042** | 1.03 | 0.98, 1.08 | 0.3 | 1.06 | 0.99–1.13 | 0.083 | 1.04 | 1.00–1.08 | 0.054 | 0.99 | 0.94, 1.04 | 0.6 | 1.02 | 0.98, 1.07 | 0.3 |
| DQB1_HED-R | 1.01 | 0.99–1.04 | 0.329 | 1.02 | 0.98, 1.06 | 0.3 | 1.00 | 0.96–1.05 | 0.835 | 1.01 | 0.98–1.04 | 0.580 | 1.03 | 0.99, 1.06 | 0.14 | 1.00 | 0.97, 1.03 | >0.9 |
| HED-MM DRB1 | 1.14 | 1.02–1.28 | **0.021** | 1.04 | 0.97, 1.11 | 0.3 | 0.98 | 0.90–1.07 | 0.682 | 1.11 | 0.99–1.24 | 0.070 | 0.95 | 0.89, 1.01 | 0.11 | 1.00 | 0.94, 1.07 | 0.9 |
| ttHED-MM DRB1 | 0.94 | 0.89–0.99 | **0.026** | — | — | — | — | — | — | 0.96 | 0.91–1.01 | 0.089 | — | — |  | — | — |  |
| ttHED-MM C | — | — | — | — | — | — | 0.85 | 0.75–0.96 | **0.011** | — | — | — | — | — | — | — | — | — |

Mismatch DQB1 Group (N=673)

|  | **RFS** | | | **Relapse** | | | **NRM** | | | **OS** | | | **aGVHD** | | | **cGVHD** | | |
| --- | --- | --- | --- | --- | --- | --- | --- | --- | --- | --- | --- | --- | --- | --- | --- | --- | --- | --- |
| **Variable** | **HR** | **95% CI** | **p** | **HR** | **95% CI** | **p** | **HR** | **95% CI** | **p** | **HR** | **95% CI** | **p** | **HR** | **95% CI** | **p** | **HR** | **95% CI** | **p** |
| **Stem-cell source** |  |  |  |  |  |  |  |  |  |  |  |  |  |  |  |  |  |  |
| BM | — | — |  | — | — |  | — | — |  | — | — |  | — | — |  | — | — |  |
| PB | 1.53 | 0.99, 2.37 | 0.057 | 1.35 | 0.79, 2.31 | 0.3 | 1.89 | 0.87, 4.13 | 0.11 | 1.49 | 0.94, 2.36 | 0.089 | 1.09 | 0.64, 1.85 | 0.8 | 1.61 | 0.97, 2.66 | 0.065 |
| Patient age (decades) | 1.05 | 0.96, 1.15 | 0.3 | 0.95 | 0.85, 1.07 | 0.4 | 1.25 | 1.06, 1.46 | 0.006 | 1.11 | 1.00, 1.23 | 0.040 | 1.02 | 0.90, 1.15 | 0.8 | 1.04 | 0.92, 1.16 | 0.5 |
| Donor age (decades) | 1.01 | 0.88, 1.15 | 0.9 | 1.01 | 0.86, 1.20 | 0.9 | 1.01 | 0.81, 1.24 | >0.9 | 0.98 | 0.85, 1.13 | 0.8 | 1.11 | 0.94, 1.32 | 0.2 | 0.97 | 0.82, 1.13 | 0.7 |
| **DRI** |  |  |  |  |  |  |  |  |  |  |  |  |  |  |  |  |  |  |
| Intermediate-Low | — | — |  | — | — |  | — | — |  | — | — |  | — | — |  | — | — |  |
| High-Very High | 0.55 | 0.44, 0.70 | <0.001 | 0.48 | 0.36, 0.64 | <0.001 | 0.71 | 0.48, 1.03 | 0.073 | 0.54 | 0.42, 0.69 | <0.001 | 1.23 | 0.88, 1.73 | 0.2 | 1.29 | 0.94, 1.77 | 0.11 |
| **T-cell depletion** |  |  |  |  |  |  |  |  |  |  |  |  |  |  |  |  |  |  |
| No | — | — |  | — | — |  | — | — |  | — | — |  | — | — |  | — | — |  |
| Yes | 0.82 | 0.60, 1.12 | 0.2 | 0.93 | 0.61, 1.41 | 0.7 | 0.69 | 0.43, 1.11 | 0.12 | 0.71 | 0.52, 0.98 | 0.037 | 0.51 | 0.36, 0.73 | <0.001 | 0.53 | 0.36, 0.77 | <0.001 |
| **Donor sex match** |  |  |  |  |  |  |  |  |  |  |  |  |  |  |  |  |  |  |
| Other | — | — |  | — | — |  | — | — |  | — | — |  | — | — |  | — | — |  |
| Male-female | 1.00 | 0.73, 1.36 | >0.9 | 0.93 | 0.61, 1.41 | 0.7 | 1.10 | 0.68, 1.79 | 0.7 | 1.00 | 0.71, 1.40 | >0.9 | 1.00 | 0.66, 1.53 | >0.9 | 1.59 | 1.13, 2.24 | 0.008 |
| **Karnofsky score** |  |  |  |  |  |  |  |  |  |  |  |  |  |  |  |  |  |  |
| ≥90 | — | — |  | — | — |  | — | — |  | — | — |  | — | — |  | — | — |  |
| <90 | 1.47 | 1.14, 1.89 | 0.003 | 1.43 | 1.03, 1.98 | 0.032 | 1.55 | 1.04, 2.31 | 0.031 | 1.67 | 1.28, 2.16 | <0.001 | 1.14 | 0.81, 1.60 | 0.5 | 0.86 | 0.61, 1.21 | 0.4 |
| **CMV -/- vs. other** |  |  |  |  |  |  |  |  |  |  |  |  |  |  |  |  |  |  |
| Other combination | — | — |  | — | — |  | — | — |  | — | — |  | — | — |  | — | — |  |
| -/- | 0.90 | 0.69, 1.18 | 0.5 | 1.12 | 0.80, 1.56 | 0.5 | 0.61 | 0.37, 1.00 | 0.052 | 0.78 | 0.58, 1.05 | 0.10 | 1.15 | 0.82, 1.63 | 0.4 | 0.95 | 0.69, 1.30 | 0.7 |
| Year transplant | 0.97 | 0.92, 1.02 | 0.2 | 0.97 | 0.91, 1.03 | 0.3 | 0.97 | 0.90, 1.05 | 0.4 | 0.95 | 0.90, 1.00 | 0.064 | 0.99 | 0.93, 1.05 | 0.7 | 0.95 | 0.90, 1.01 | 0.081 |
| **Total body irradiation** |  |  |  |  |  |  |  |  |  |  |  |  |  |  |  |  |  |  |
| No | — | — |  | — | — |  | — | — |  | — | — |  | — | — |  | — | — |  |
| Yes | 0.90 | 0.68, 1.20 | 0.5 | 0.82 | 0.56, 1.18 | 0.3 | 1.05 | 0.67, 1.64 | 0.8 | 0.90 | 0.66, 1.22 | 0.5 | 1.19 | 0.83, 1.70 | 0.3 | 0.71 | 0.50, 1.00 | 0.050 |
| **Conditioning intensity** |  |  |  |  |  |  |  |  |  |  |  |  |  |  |  |  |  |  |
| Standard | — | — |  | — | — |  | — | — |  | — | — |  | — | — |  | — | — |  |
| Reduced | 1.00 | 0.77, 1.30 | >0.9 | 1.14 | 0.81, 1.61 | 0.4 | 0.81 | 0.53, 1.23 | 0.3 | 0.96 | 0.72, 1.27 | 0.8 | 0.82 | 0.57, 1.17 | 0.3 | 0.86 | 0.63, 1.17 | 0.3 |
| **GVHD prophylaxis** |  |  |  |  |  |  |  |  |  |  |  |  |  |  |  |  |  |  |
| CSA-MTX | — | — |  | — | — |  | — | — |  | — | — |  | — | — |  | — | — |  |
| CSA-MMF | 1.14 | 0.85, 1.53 | 0.4 | 1.11 | 0.75, 1.62 | 0.6 | 1.15 | 0.72, 1.83 | 0.6 | 1.03 | 0.75, 1.42 | 0.8 | 1.07 | 0.72, 1.60 | 0.7 | 1.24 | 0.88, 1.76 | 0.2 |
| Tac-based | 1.19 | 0.84, 1.69 | 0.3 | 1.40 | 0.91, 2.16 | 0.13 | 0.86 | 0.47, 1.58 | 0.6 | 1.14 | 0.79, 1.64 | 0.5 | 0.96 | 0.59, 1.55 | 0.9 | 1.01 | 0.65, 1.57 | >0.9 |
| Other | 1.39 | 1.00, 1.93 | 0.049 | 1.35 | 0.88, 2.08 | 0.2 | 1.42 | 0.85, 2.38 | 0.2 | 1.31 | 0.93, 1.86 | 0.13 | 1.18 | 0.75, 1.86 | 0.5 | 1.22 | 0.83, 1.81 | 0.3 |
| A_HED-R | 1.01 | 0.98, 1.04 | 0.7 | 1.00 | 0.96, 1.03 | 0.8 | 1.02 | 0.97, 1.07 | 0.4 | 1.02 | 0.99, 1.05 | 0.3 | 1.03 | 0.99, 1.07 | 0.2 | 1.00 | 0.96, 1.03 | >0.9 |
| B_HED-R | 1.00 | 0.97, 1.04 | >0.9 | 1.02 | 0.98, 1.07 | 0.4 | 0.97 | 0.91, 1.03 | 0.3 | 1.00 | 0.96, 1.04 | >0.9 | 1.02 | 0.97, 1.07 | 0.5 | 0.98 | 0.94, 1.03 | 0.4 |
| C_HED-R | 1.05 | 0.99, 1.10 | 0.088 | 1.04 | 0.98, 1.12 | 0.2 | 1.05 | 0.96, 1.14 | 0.3 | 1.06 | 1.00, 1.12 | **0.045** | 1.02 | 0.95, 1.10 | 0.6 | 0.99 | 0.93, 1.05 | 0.6 |
| DRB1_HED-R | 0.98 | 0.96, 1.00 | 0.069 | 0.97 | 0.94, 1.00 | 0.067 | 0.99 | 0.95, 1.03 | 0.5 | 0.98 | 0.95, 1.00 | 0.054 | 1.00 | 0.97, 1.03 | >0.9 | 1.00 | 0.98, 1.03 | 0.8 |
| DQB1_HED-R | 1.00 | 0.98, 1.02 | 0.8 | 1.00 | 0.97, 1.03 | >0.9 | 1.01 | 0.98, 1.04 | 0.6 | 1.02 | 1.00, 1.04 | 0.12 | 1.01 | 0.99, 1.04 | 0.4 | 1.00 | 0.98, 1.03 | 0.8 |
| HED-MM DQB1 | 0.99 | 0.95, 1.02 | 0.5 | 0.97 | 0.93, 1.01 | 0.14 | 1.02 | 0.96, 1.07 | 0.5 | 0.99 | 0.96, 1.03 | 0.7 | 1.01 | 0.96, 1.05 | 0.7 | 0.99 | 0.95, 1.03 | 0.5 |

*HR = Hazard Ratio; CI = Confidence Interval; tt = time-dependent analysis (time interaction). Reference categories are indicated by '—'. Statistically significant results for HED metrics are highlighted (p<0.05)*

##### **Table S4: DPB1 mismatches across other locus-mismatches**

| **Mismatched locus** | **DP mismatch** |
| --- | --- |
| **N** | N = 1389 |
| **MM-A** | 448 (32%) |
| **MM-B** | 217 (16%) |
| **MM-C** | 352 (25%) |
| **MM-DRB1** | 126 (9.1%) |
| **MM-DQB1** | 246 (18%) |

##### **Table S5: DPB1 focused analysis**

| **Outcome** | **DPB1 HED-R HR (95% CI)** | **p-value** | **DPB1 HED-MM HR (95% CI)** | **p-value** |
| --- | --- | --- | --- | --- |
| **Overall Survival** | 0.98 (0.95–1.01) | 0.14 | 1.00 (0.98–1.01) | 0.50 |
| **Relapse-Free Survival** | 0.99 (0.96–1.01) | 0.30 | 1.00 (0.98–1.01) | 0.50 |
| **Relapse** | 0.98 (0.95–1.02) | 0.40 | 1.00 (0.98–1.02) | >0.90 |
| **Non-Relapse Mortality** | 0.99 (0.95–1.03) | 0.50 | 0.99 (0.97–1.01) | 0.40 |
| **aGvHD II–IV** | 0.96 (0.92–1.01) | 0.13 | 0.99 (0.98–1.01) | 0.50 |
| **cGvHD** | 0.99 (0.96–1.03) | 0.70 | 0.99 (0.97–1.01) | 0.40 |
| (OS N=1161 , RFS, RI, NRM N=1144, aGvHD N=1129 , cGvHD N=1133) Each model is built on the subgroup DPB1 mismatch and adjusted for the following clinical variables: for stem cell source, recipient and donor age, disease risk index, T-cell depletion, donor–recipient sex mismatch, Karnofsky score, CMV serostatus, year of transplant, conditioning intensity, total body irradiation, and GvHD prophylaxis, as well as for locus-specific HED-R. | | | | |
